# An integrated analysis of contact tracing and genomics to assess the efficacy of travel restrictions on SARS-CoV-2 introduction and transmission in England from June to September, 2020

**DOI:** 10.1101/2021.03.15.21253590

**Authors:** Dinesh Aggarwal, Andrew J. Page, Ulf Schaefer, George M. Savva, Richard Myers, Erik Volz, Nicholas Ellaby, Steven Platt, Natalie Groves, Eileen Gallaghar, Niamh M. Tumelty, Thanh Le Viet, Gareth J. Hughes, Cong Chen, Charlie Turner, Sophie Logan, Abbie Harrison, The COVID-19 Genomics UK (COG-UK) Consortium, Sharon J. Peacock, Meera Chand, Ewan M. Harrison

## Abstract

**Background:** Mitigation of SARS-CoV-2 transmission from international travel is a priority. Travellers from countries with travel restrictions (closed travel-corridors) were required to quarantine for 14 days over Summer 2020 in England. We describe the genomic epidemiology of travel-related cases in England and evaluate the effectiveness of this travel policy.

**Methods:** Between 27/05/2020 and 13/09/2020, probable travel-related SARS-CoV-2 cases and their contacts were identified and combined with UK SARS-CoV-2 sequencing data. The epidemiology and demographics of cases was identified, and the number of contacts per case modelled using negative binomial regression to estimate the effect of travel restriction, and any variation by age, sex and calendar date. Unique travel-related SARS-CoV-2 genomes in the COG-UK dataset were identified to estimate the effect travel restrictions on cluster size generated from these. The Polecat Clustering Tool was used to identify a travel-related SARS-CoV-2 cluster of infection.

**Findings:** 4,207 travel-related SARS-CoV-2 cases are identified. 51.2% (2155/4207) of cases reported travel to one of three countries; 21.0% (882) Greece, 16.3% (685) Croatia and 14.0% (589) Spain. Median number of contacts per case was 3 (IQR 1-5), and greatest for the 16-20 age-group (9.0, 95% C.I.=5.6-14.5), which saw the largest attenuation by travel restriction. Travel restriction was associated with a 40% (rate ratio=0.60, 95% C.I.=0.37-0.95) lower rate of contacts. 827/4207 (19.7%) of cases had high-quality SARS-CoV-2 genomes available. Fewer genomically-linked cases were observed for index cases related to countries with travel restrictions compared to cases from non-travel restriction countries (rate ratio=0.17, 95% C.I.=0.05-0.52). A large travel-related cluster dispersed across England is identified through genomics, confirmed with contact-tracing data.

**Interpretation:** This study demonstrates the efficacy of travel restriction policy in reducing the onward transmission of imported cases.

**Funding:** Wellcome Trust, Biotechnology and Biological Sciences Research Council, UK Research & Innovation, National Institute of Health Research, Wellcome Sanger Institute.

**RESEARCH IN CONTEXT:** *Evidence before this study:* We searched PubMed, medRxiv, bioRxiv, Web of Science and Scopus for the terms (COVID-19 OR SARS-COV-2) AND (imported or importation) AND (sequenc* OR genom* or WGS). We filtered the 55 articles identified through this search and rejected any that did not undertake SARS-CoV-2 sequencing as part of an epidemiological investigation for importation into a different country. The remaining 20 papers were reviewed in greater detail to understand the patterns of importation and the methods used in each case.

*Added value of this study:* This is the first published study on importations of SARS-CoV-2 into England using genomics. Plessis et al., (2021) used a predictive model to infer the number of importations in to the UK from all SARS-CoV-2 genomes generated before 26th June 2020. The current study assesses the period 27/05/2020 to 13/09/2020 and presents findings of case-reported travel linked to genomic data. Two unpublished reports exist for Wales and Scotland, although only examine a comparatively small number of importations.

*Implications of all the available evidence:* This large-scale study has a number of findings that are pertinent to public health and of global significance, not available from prior evidence to our knowledge. The study demonstrates travel restrictions, through the implementation of ‘travel-corridors’, are effective in reducing the number of contacts per case based on observational data. Age has a significant effect on the number of contacts and this can be mitigated with travel restrictions. Analysis of divergent clusters indicates travel restrictions can reduce the number of onwards cases following a travel-associated case. Analysis of divergent clusters can allow for importations to be identified from genomics, as subsequently evidenced by cluster characteristics derived from contact tracing. The majority of importations of SARS-CoV-2 in England over Summer 2020 were from coastal European countries. The highest number of cases and onward contacts were from Greece, which was largely exempt from self-isolation requirements (bar some islands in September at the end of the study period). Systematic monitoring of imported SARS-CoV-2 cases would help refine implementation of travel restrictions. Finally, along with multiple studies, this study highlights the use of genomics to monitor and track importations of SARS-CoV-2 mutations of interest; this will be of particular use as the repertoire of clinically relevant SARS-CoV-2 variants expand over time and globally.

## Introduction

A new coronavirus related disease (COVID-19) was first reported in Wuhan, China (2) in December 2019, with the causative virus identified as a novel coronavirus SARS-CoV-2 (3). Since then, SARS-CoV-2 has been imported into virtually every country and region in the world. Understanding and tracking the sources of importations can give important information for policy makers, and for managing the pandemic, by informing policies aimed at reducing the further spread of virus.

Public health measures can help mitigate and suppress the spread of the virus, but the threat of importations will remain. The available brakes on imported SARS-CoV-2 cases include travel bans, quarantine measures, and testing of returning travellers. These can apply to all countries or targeted to high-risk countries, for variable durations, and with variable degree of enforcement. In England, travel restrictions were assigned on a country by country basis from 6 July 2020 entailing the use of ‘travel-corridors’; travellers returning from countries that were on the travel restrictions list (with ‘closed travel-corridors’) (4) were required to quarantine for 14 days (reduced to 10 days on 15/12/20), or from the 15^th^ December 2020, choose to quarantine for 5 days and then pay for a SARS-CoV-2 diagnostic test (Figure 1). This policy aims to limit onwards transmission of SARS-CoV-2, and as a secondary outcome possibly deter travel to those countries. Upon identification of an imported case, contact tracing and quarantine/self-isolation measures can limit onwards transmission. The CORSAIR study reported that 18.2% of individuals adhered to general SARS-CoV-2 self-isolation guidance recommended by Public Health England in the UK (5). The PHE Isolation Assurance Service however have identified up to 97% self-reported compliance with travel-specific self-isolation guidance (6). These data do not include countries exempt from quarantine, contact-tracing data or link to genomic data to evaluate travel-related clusters.

**Figure 1a:**
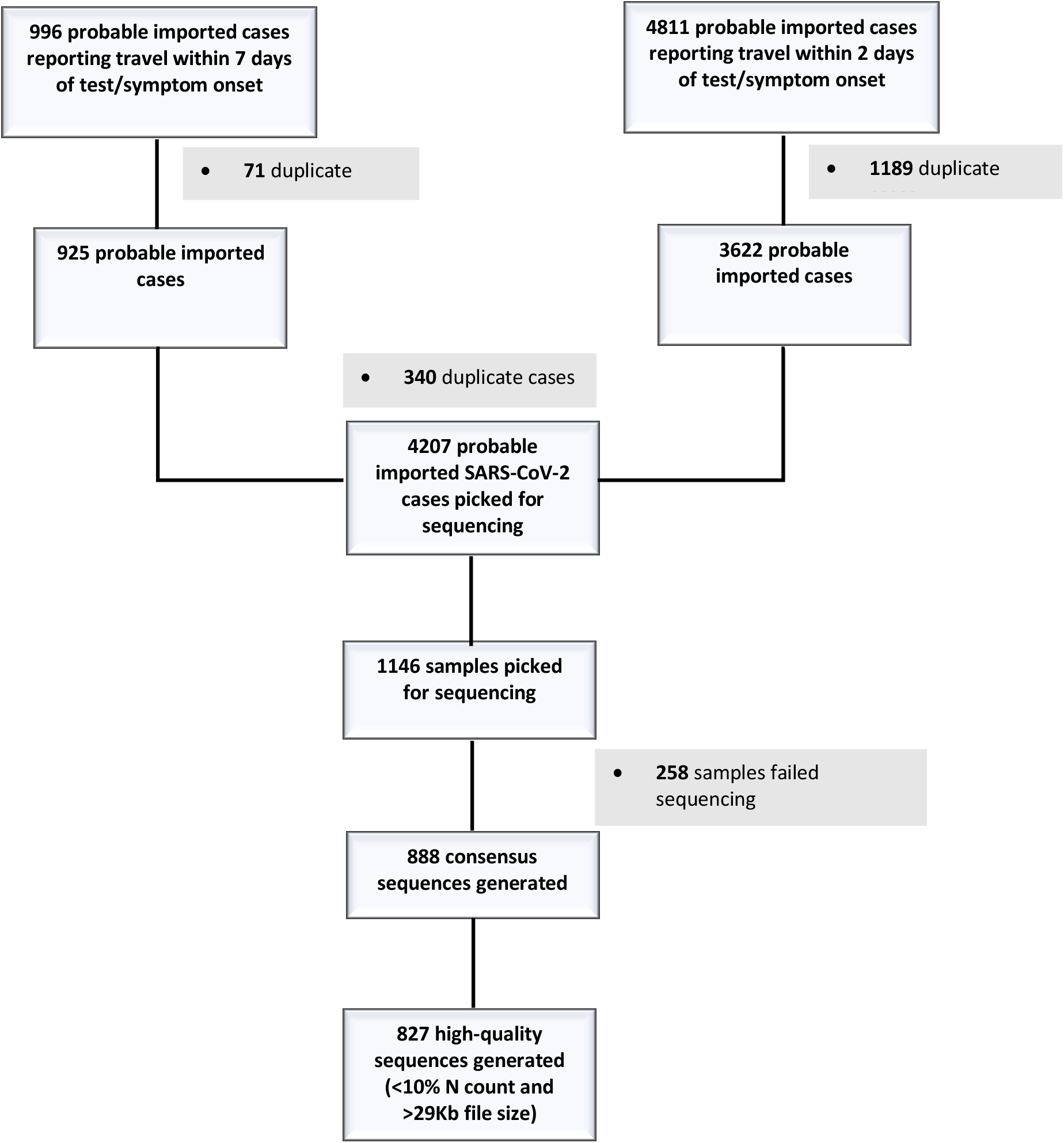
Flow diagram of travel-related case ascertainment from Test and Trace data and subsequent genome availability. Cases were defined as ‘highly probable’ and ‘probable’. ‘Highly probable’ travel-related cases were defined as individuals who reported international travel as an activity in the two days before symptom onset/testing. On 12/08/2020 the additional facility to report international travel in the seven days prior to symptom onset/testing became available, and also included in this study and defined as ‘probable’ travel-related cases.

**Figure 1b:**
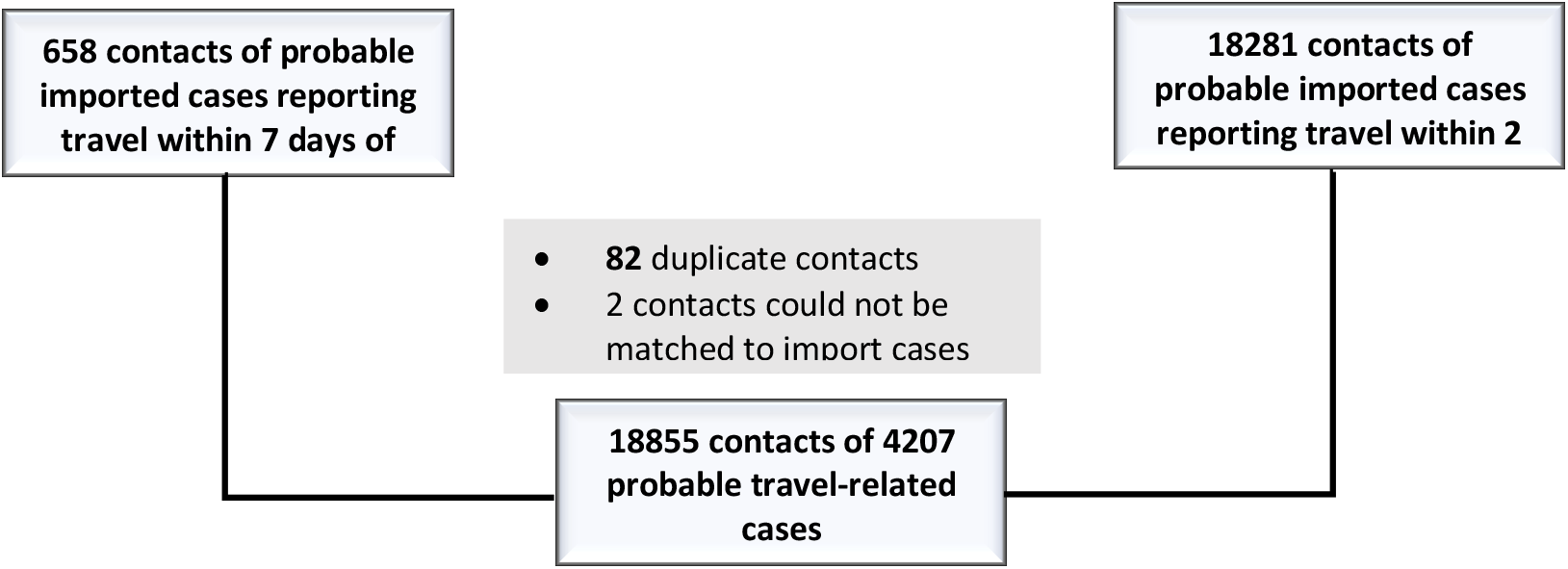
Flow diagram relaying contacts ascertained of cases from Test and Trace data.

**Figure 1c:**
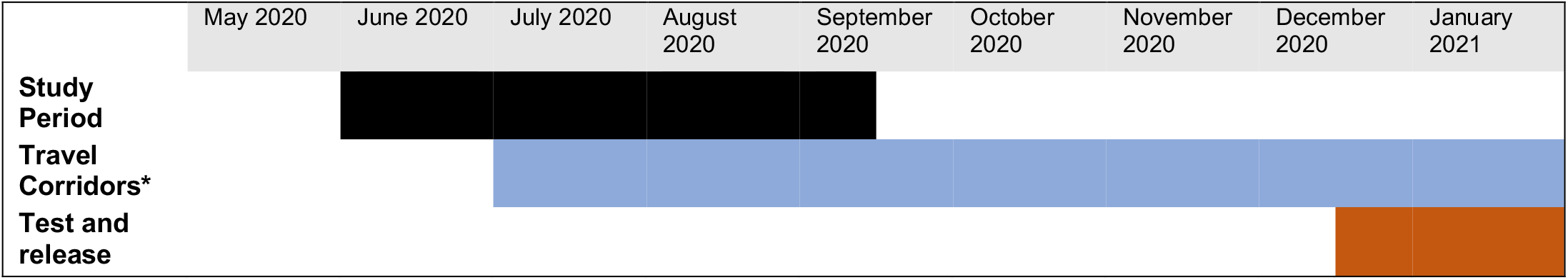
Timeline of study period (27/05/2020 to 13/09/2020) and associated policy changes on travel introduced in England. Travel restrictions were assigned on a country by country basis from 6 July 2020. Travellers returning from countries that were on the travel restrictions list (4) were required to self-isolate for 14 days (*reduced to 10 days on 15/12/20), or from the 15^th^ December 2020, choose to self-isolate for 5 days and then pay for a SARS-CoV-2 diagnostic test (test and release)

Studies from numerous countries have used genome sequencing to complement epidemiological investigations in order to characterise importations of SARS-CoV-2 (Supplementary Table 1). Primarily these are in-depth case reports on small datasets but demonstrate the utility of genomics combined with contact tracing. Genomic sequencing of returning travellers was useful in allowing for the first case of reinfection of SARS-CoV-2 in the world to be identified in Hong Kong (7) and identify a new variant of SARS-CoV-2 (B.1.177/20A.EU1, variant A222V) (8).

This study combines contact-tracing data from National Health Service (NHS) Test and Trace (T&T) for probable importation cases with genomic data made available through the COVID-19 Genomics UK (COG-UK) consortium (9), which receives samples from NHS hospital diagnostic labs and mass community testing labs (UK Lighthouse labs network) across the UK. We aimed to characterise the known imported cases and the effectiveness of travel restrictions on onwards transmission.

A total of 4,207 SARS-CoV-2 positive importation cases were analysed, along with 18,856 contacts, of which 888 sequenced genomes were available for comparison to all UK genomic data (131,387 sequences from the UK and in the COG-UK dataset by 5 December 2020). The number of contacts reported by a case was used as an indicator of adherence to quarantining.

## Methods and materials

### Contact tracing and case identification

Contact-tracing data was obtained from T&T. All cases and contacts had a field for demographic data, but this was not always reported (Table 2 and Supplementary Table 3). ‘Highly probable’ travel-related cases were defined as individuals who reported international travel as an activity in the two days before symptom onset/testing. On 12/08/2020 the additional facility to report international travel in the seven days prior to symptom onset/testing became available, and also included in this study and defined as ‘probable’ travel-related cases.

**Table 1:** The top 20 countries reported as the travel destination for importations of SARS-CoV-2 into England and the associated number of samples sequenced from travel-related cases.

| Country | Cases |  | Sequenced samples from cases (passed QC) |  | Percentage of cases sequenced by country of travel |
| --- | --- | --- | --- | --- | --- |
|  | N | % | N | % |  |
|  |  |  |  |  | 18.8% |
| Greece | 882 | 21.0% | 166 | 20.1% | 23.6% |
| Croatia | 685 | 16.3% | 162 | 19.6% | 18.0% |
| Spain | 589 | 14.0% | 106 | 12.8% | 20.2% |
| Unknown | 282 | 6.7% | 57 | 6.9% | 23.3% |
| France | 223 | 5.3% | 52 | 6.3% | 11.2% |
| Turkey | 187 | 4.4% | 21 | 2.5% | 18.0% |
| Portugal | 111 | 2.6% | 20 | 2.4% | 15.2% |
| Malta | 99 | 2.4% | 15 | 1.8% | 22.6% |
| Italy | 93 | 2.2% | 21 | 2.5% | 16.5% |
| Poland | 85 | 2.0% | 14 | 1.7% | 16.7% |
| Romania | 78 | 1.9% | 13 | 1.6% | 15.4% |
| Czech Republic | 65 | 1.5% | 10 | 1.2% | 19.7% |
| Albania | 61 | 1.4% | 12 | 1.5% | 27.9% |
| Hungary | 61 | 1.4% | 17 | 2.1% | 24.6% |
| India | 57 | 1.4% | 14 | 1.7% | 21.8% |
| Pakistan | 55 | 1.3% | 12 | 1.5% | 13.2% |
| Netherlands | 38 | 0.9% | 5 | 0.6% | 20.7% |
| Germany | 29 | 0.7% | 6 | 0.7% | 25.0% |
| Switzerland | 28 | 0.7% | 7 | 0.8% | 12.5% |
| Kosovo | 24 | 0.6% | 3 | 0.4% | 18.8% |
| **Total cases** | 4207 |  | 827 |  | 19.7% |

**Table 2:** Contacts per case related to Sex, Age, Ethnic Group, Region of residence and reported Travel Destination.

| Demographic | Cases | Total contacts of cases | contacts reported per case |
| --- | --- | --- | --- |
| **Sex** |  |  |  |
| Male | 2193 | 9835 | 4.5 |
| Female | 1933 | 8578 | 4.4 |
| Unknown | 82 | 224 | 3.1 |
| **Age** |  |  |  |
| 0-5 | 51 | 183 | 3.6 |
| 6-10 | 45 | 124 | 2.8 |
| 11-15 | 75 | 321 | 4.3 |
| 16-20 | 1086 | 7473 | 6.9 |
| 21-25 | 843 | 3536 | 4.2 |
| 26-30 | 685 | 2091 | 3.1 |
| 31-35 | 413 | 1312 | 3.2 |
| 36-40 | 278 | 939 | 3.4 |
| 41-45 | 185 | 566 | 3.1 |
| 46-50 | 169 | 723 | 4.3 |
| 51-55 | 130 | 469 | 3.6 |
| 56-60 | 121 | 434 | 3.6 |
| 61-65 | 57 | 229 | 4.0 |
| 66-70 | 30 | 116 | 3.9 |
| 71-75 | 20 | 63 | 3.2 |
| 76-80 | 7 | 16 | 2.3 |
| 81-85 | 7 | 39 | 5.6 |
| 86-90 | 3 | 3 | 1.0 |
| 91-95 | 0 | 0 | NA |
| Unknown | 2 |  |  |
| **Ethnic group** |  |  |  |
| **White** |  |  |  |
| English/Welsh/Scottish/Northern Irish/British | 2509 | 12745 | 5.1 |
| Irish | 35 | 121 | 3.5 |
| Gypsy or Irish Traveller | 0 | 0 | NA |
| Any other White background | 583 | 1755 | 3.0 |
| **Mixed/Multiple ethnic groups** |  |  |  |
| White and Black Caribbean | 42 | 284 | 6.8 |
| White and Black African | 25 | 108 | 4.3 |
| White and Asian | 49 | 216 | 4.4 |
| Any other Mixed/Multiple ethnic background | 44 | 147 | 3.3 |
| **Asian/Asian British** |  |  |  |
| Indian | 103 | 481 | 4.7 |
| Pakistani | 79 | 306 | 3.9 |
| Bangladeshi | 27 | 82 | 3.0 |
| Chinese | 6 | 2 | 0.3 |
| Any other Asian background | 62 | 199 | 3.2 |
| **Black/ African/Caribbean/Black British** |  |  |  |
| African | 58 | 152 | 2.6 |
| Caribbean | 12 | 36 | 3.0 |
| Any other Black/African/Caribbean background | 12 | 28 | 2.3 |
| **Other ethnic group** |  |  |  |
| Arab | 0 | 0 | NA |
| Any other ethnic group | 110 | 367 | 3.3 |
| **Other** |  |  |  |
| Prefer not to say | 65 | 135 | 2.1 |
| Unknown | 386 | 1473 | 3.8 |
| **Region** |  |  |  |
| London | 1205 | 4275 | 3.5 |
| South East | 622 | 3211 | 5.2 |
| North West | 584 | 2323 | 4.0 |
| East of England | 395 | 2079 | 5.3 |
| South West | 328 | 1960 | 6.0 |
| Yorkshire and Humber | 327 | 1411 | 4.3 |
| West Midlands | 299 | 1259 | 4.2 |
| East Midlands | 251 | 1351 | 5.4 |
| North East | 161 | 660 | 4.1 |
| Not stated | 35 | 108 | 3.1 |
| **Country** |  |  |  |
| Greece | 882 | 5587 | 6.3 |
| Croatia | 685 | 3913 | 5.7 |
| Spain | 589 | 1521 | 2.6 |
| Unknown | 282 | 988 | 3.5 |
| France | 223 | 815 | 3.7 |
| Turkey | 187 | 702 | 3.8 |
| Portugal | 111 | 439 | 4.0 |
| Malta | 99 | 492 | 5.0 |
| Italy | 93 | 390 | 4.2 |
| Poland | 85 | 417 | 4.9 |
| Romania | 78 | 189 | 2.4 |
| Hungary | 67 | 137 | 2.0 |
| Czech Republic | 66 | 239 | 3.6 |
| Albania | 61 | 140 | 2.3 |
| India | 57 | 223 | 3.9 |
| Pakistan | 55 | 269 | 4.9 |
| Netherlands | 38 | 166 | 4.4 |
| Germany | 29 | 101 | 3.5 |
| Switzerland | 28 | 123 | 4.4 |
| Kosovo | 24 | 64 | 2.7 |

Cases were asked to provide details of all contacts for activities in the 2 days prior to onset/testing up to completing the system which were gathered. If any contacts become cases they would then also be included in T&T data as a case separately, but if they did not report direct travel themselves, then they would not meet the definition for a travel-associated case.

### Case identification from T&T data

Data included free-text destination city or country. A freetext country and city search with a custom python script on travel-related T&T was used to identify destination country. Results and remaining entries were manually checked and corrected (see Supplementary methods for more details).

### Clinical samples, Genome sequencing and Quality Control

Clinical samples were collected passively as part of national SARS-CoV-2 testing. This included both community testing through lighthouse labs and testing through hospital diagnostic labs. Samples were sequenced at one of seventeen COG-UK sequencing sites (Figure 1). The samples were prepared for sequencing using either the ARTIC (10) or veSeq (11) protocols, and were sequenced using Illumina or Oxford Nanopore platforms. All samples were uploaded to and processed through COVID-CLIMB pipelines (12,13). Genomes were aligned to the Wuhan Hu-1 reference genome (MN908947.3). Genomes which contained more than 10% missing data were excluded from further analysis to ensure high quality phylogenetic analysis.

### Lineages and minor variants

Global and UK Lineages (14) were assigned to each genome using Pangolin (https://github.com/cov-lineages/pangolin) with analysis performed on COVID-CLIMB (13). Minor variants were pre-defined within the COG-UK database using type_variants (https://github.com/cov-ert/type_variants).

### Identification of extinct and unique genomes

The 827 high-quality travel-related genomes were compared to the COG-UK dataset on 16/10/2020. Genomes were only compared to other genomes with the same UK lineage assigned by COG-UK, since we assume that no relatedness relevant to transmission exists between genomes of different UK lineages. A ‘unique’ genome in the community was deemed to be one that was known to be from a travel-related case and either: (1) A UK lineage that had not been sampled in the previous 4 weeks in the UK, (2) >3 SNPs distance to the closest relative in the COG-UK dataset.

Within the same UK lineage we identified those genomes sampled within 4 weeks prior to the genome of interest. We determined the minimum SNP distance between the sequence of interest and these genomes. ‘Unique’ genomes were compared to sequences that were generated in the COG-UK dataset within 2 and 4 weeks after their sampling date, to identify samples with the same UK lineage and within 2 SNPs. These would represent onward transmission or further introductions of similar genomes. The analysis was run with an in-house custom Python script developed by US and RM. Further detail in supplementary methods.

### Identification of a travel-related SARS-CoV-2 cluster

We used the Polecat clustering tool (https://cog-uk.github.io/polecat) to systematically identify outliers in COG-UK genomic dataset and link to contact-tracing data.

### Statistical analysis

All models were estimated using the glmmTMB package (version 1.0.1) (15) with marginal means and effects calculated using the emmeans package (1.5.2-1) (16) for R (version 3.5.1) (17). Figures were generated using R (version 4.0.2) and Microsoft Excel (version 1908). The number of contacts per case was modelled using negative binomial regression analysis, to estimate the effect of travel restriction, and whether this varied by age-group, sex of the index case and calendar date. Travel destination and ethnic group were included as covariates (as random effects). A similar approach was taken when estimating the effect of travel restriction on genomic cluster size.

## Results

From 17/03/20 – 04/07/20 the Foreign & Commonwealth Office advised against all non-essential travel worldwide (18). From the 04/07/20 – 01/02/21 travel corridors to countries deemed to be low risk for COVID-19 disease (subject to assessment and change) were established in which returning travellers were no-longer required to quarantine. Persons returning from countries outside this list (except for exemptions e.g. specific employment) were required to quarantine. We sought to both gauge the impact of this policy and to attempt to quantify the numbers of onward transmissions using genomic epidemiology.

Between 27/05/2020 and 13/09/2020, using contact-tracing data for cases who have tested positive for SARS-CoV-2, we identified 4,207 travel-related cases (Figure 1). Supplementary tables 2 and 3 show the case characteristics.

Travel to European countries accounted for 85.9% (3612/4207) of cases, of which 51.2% (2155/4207) had visited one of Greece (21.0%, 882/4207), Croatia (16.3%, 685/4207) and Spain (14.0%,589/4207) (Figure 2 and Table 2). For 284 cases the country of travel was unclear or unknown. Travel restrictions were first eased on 03/07/2020; 2.9% of travel-related cases were recorded before this date. For the countries associated with the highest numbers of imports, the duration of the peak of imported cases differs, with variable association with changes in travel restriction policy (Figure 3). Geographically variations in imported cases across England were apparent, with the greatest number in Greater London (28.6%, 1205/4207) (Figure 2, Supplementary Figures 1 and 2, and Supplementary Table 3).

**Figure 2a:**
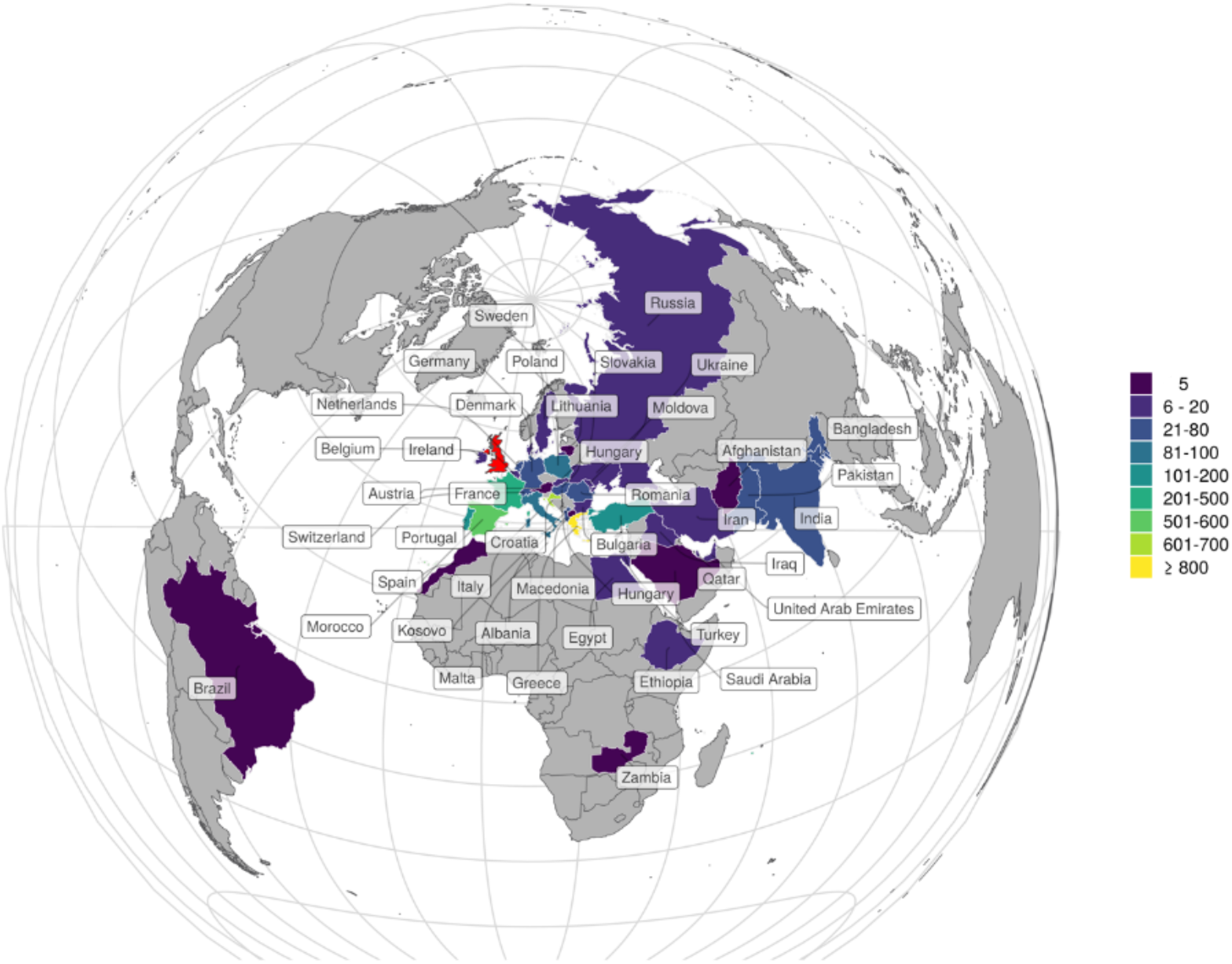
Countries where importations originated. Countries with less than 5 importations were excluded for confidentiality reasons.

**Figure 2b:**
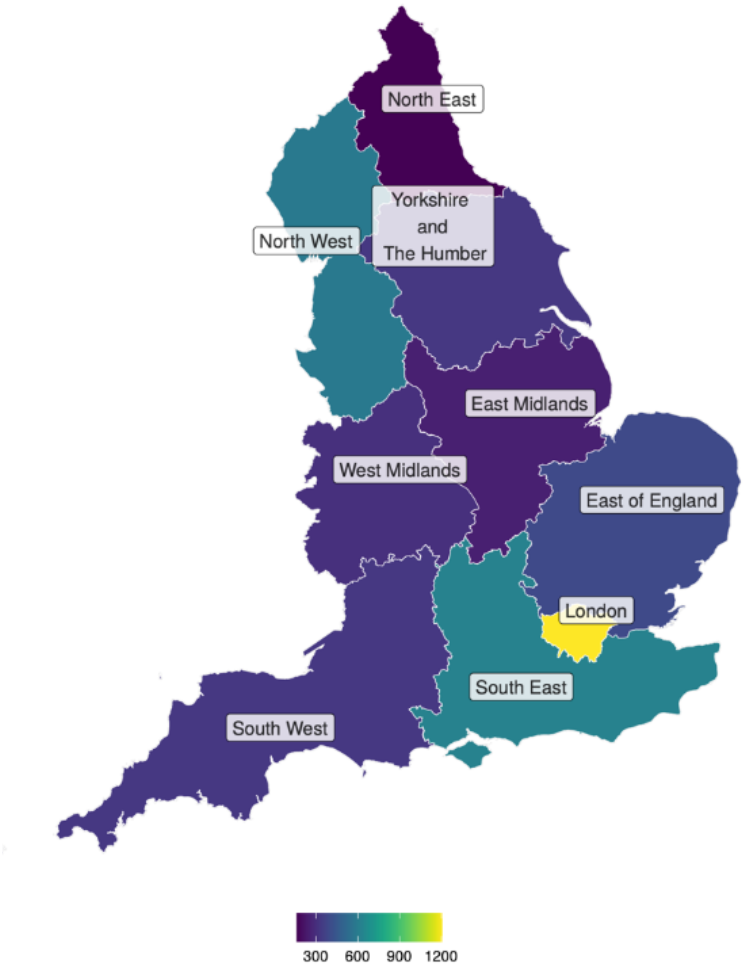
Destinations of imported cases within England. Areas with less than 3 cases have been excluded.

**Figure 3:**
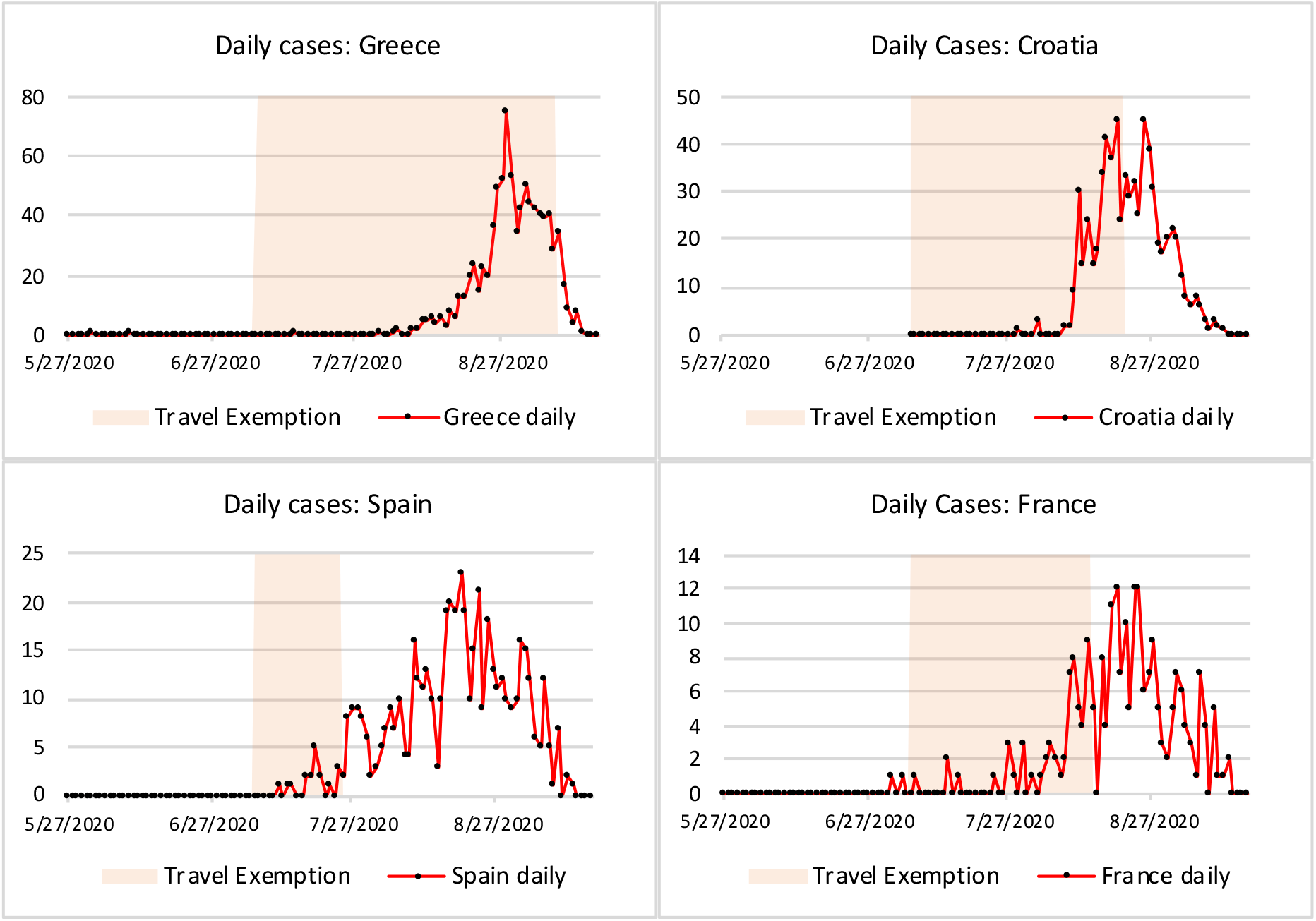
Frequency of importations over time for the top 4 most common countries of travel reported by individuals testing positive for SARS-CoV-2 during the study period. SARS-CoV-2 case numbers in returning travellers by the four most popular countries of travel reported by cases representing 2379/4207 (56.5%) of known travel-related cases. The shaded areas represent the period of time when the countries did not have restrictive travel guidance in place.

The median number of reported contacts per travel-associated case was 3 (IQR 1-5), with a maximum of 172. Overall, travel restriction reduced the number of contacts per case by 40% (rate ratio (R.R.)=0.60, 95% C.I.=0.37-0.95). The mean number of contacts (adjusting and averaging over all over covariates) was 5.85 when no travel restriction was in place and 3.50 when there was. The effect of travel restriction varied significantly with age-group and over time (Supplementary Table 4 and Figure 4). The number of contacts per case was greatest for the 16-20 age-group without travel restriction with a marginal mean of 9.0 (95% C.I.=5.6-14.5) but with restriction reduced to 4.7 (95% C.I.=3.9-5.7), and similar to other age-groups. After adjusting for all other covariates the numbers of contacts per imported case was roughly half in September compared to May, June and July, whether or not a travel restriction was in place.

**Figure 4:**
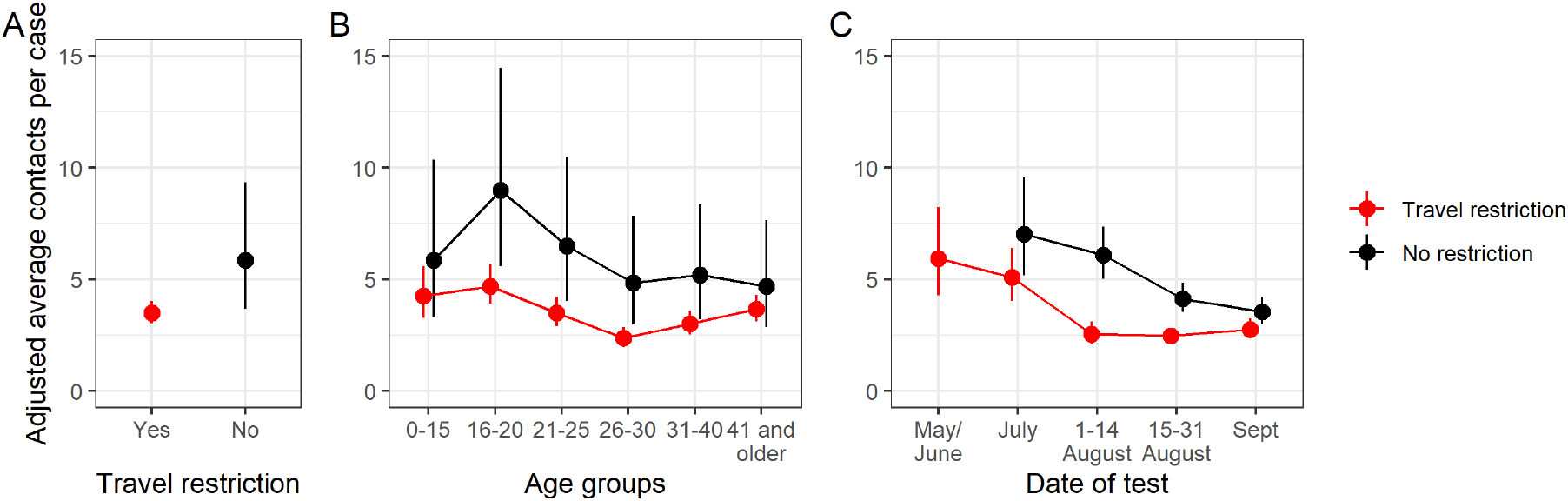
The effect of travel restriction on contacts per imported case of SARS-CoV-2. Estimated marginal mean number of contacts per imported case (a) overall, (b) by age-group and (c) by date of test comparing countries with travel restriction guidance (closed ‘travel-corridors’) in place and those without (open ‘travel-corridors). All estimates are provided with 95% confidence intervals.

### Transmission patterns identified by analysis of traveller SARS-CoV-2 genomes

We next sought to quantify onward transmission from an imported case using genomics. High-quality sequencing data was available for 827/4207 (19.7%) of cases (Figure 1) and demographics of the sequenced cases was broadly similar to the entire travel-related cohort (Supplementary Table 3).

186/827 (22.4%) imported cases had viral lineages that were sufficiently unique in the COG-UK dataset to monitor onward spread. Of these, 146/186 isolates had not been sampled in the entire UK dataset in the 4 weeks prior and 40/186 isolates were >3 SNPs to their closest matching sequence in the UK dataset.

To compare the effect of travel restrictions on the subsequent spread of likely imported cases (excluding 18/186 cases before 14/07/2020 to ensure the dates of cases with and without a travel restriction overlapped), the entire COG-UK dataset was interrogated to identify isolates within 2 SNPs of these distinct imported cases during the period 0-2 and 0-4 weeks following the importation case. The number of subsequent cases detected during the four weeks since the unique index case increased from a mean of 1.2 new cases where a travel restriction was in place to 11.3 cases where there was not. The proportions of cases leading to a subsequent newly detected case (e.g. likely transmission), and the number of new cases where at least one is detected are shown in Figure 5. Overall, 56/168 of genomes from cases that were genetically unique were detected in subsequent cases (up to four weeks later). Among cases diagnosed after returning from a country where a travel restriction was in place, 25% of (20/81) were detected in later cases (up to four weeks later), rising to 41% (29/71) when cases were imported from a country without a travel restriction. Destination country for 16 index cases was unknown. There was a high variation in the number of subsequent cases matching each genome (range 1 to 210, IQR 0-4) with a small number of imported cases corresponding to large numbers of subsequent cases (Figure 5).

**Figure 5:**
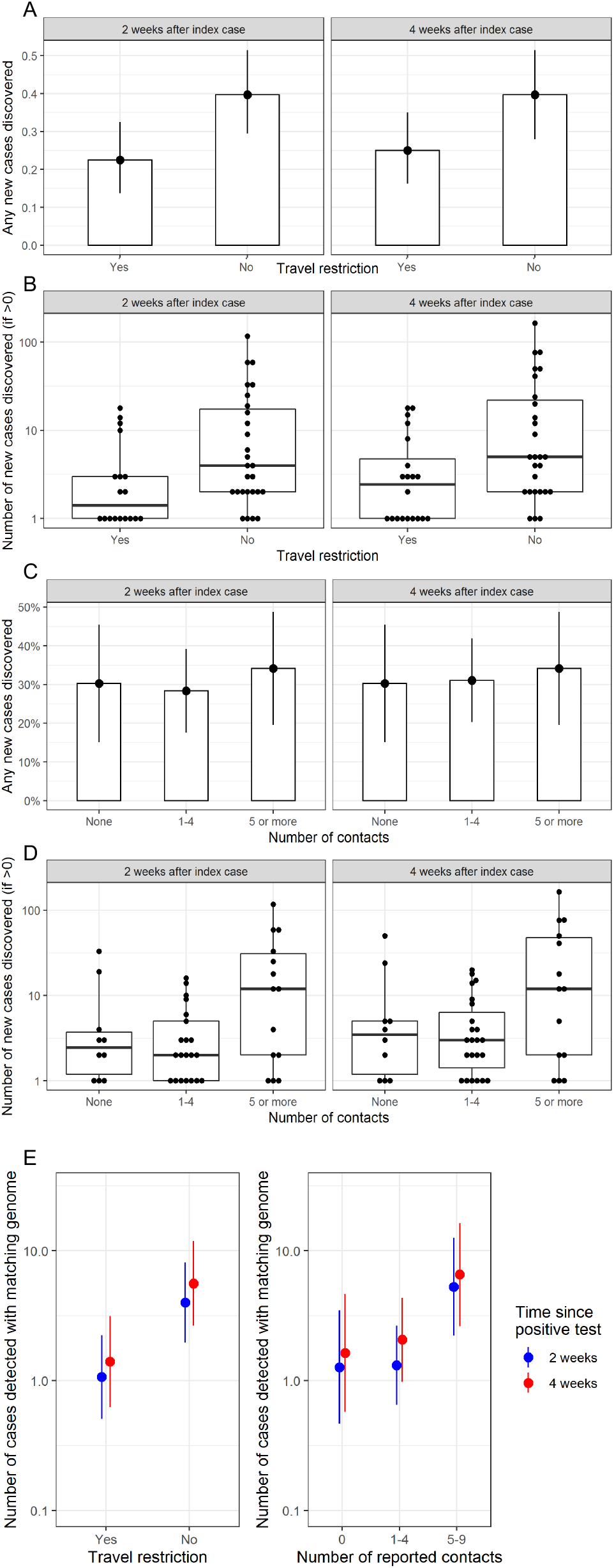
The effect of travel restrictions on the subsequent spread of likely imported cases as determined by genomics. Panel A: The proportion of imported cases with any matching genome detected over the two or four weeks following index test result. Panel B: The number of genomes matching the index case, with zeros excluded. Panel A and B compare countries with travel restriction guidance (closed ‘travel-corridors’) in place and those without (open ‘travel-corridors). Panel C: The proportion of imported cases with any matching genome detected over the two of four weeks following index test result. Panel D: The number of genomes matching the index case, with zeros excluded. Panel E: Estimated marginal mean number of genomes detected after 2 weeks or 4 weeks matching an index genome, stratified by travel restriction and stratified by number of contacts. In all panels, boxes correspond to median and interquartile range, and error bars correspond to 95% confidence intervals.

There was some evidence that imported cases with higher numbers of contacts gave rise to more cases in the subsequent month (Figure 5). Although the number of cases with any subsequent matching genome was not affected by the number of contacts, the average number of subsequent cases detected was substantially higher when the index case had five or more contacts (mean=10.8) compared to none (mean=2.8) or 1 to 4 contacts (mean=1.7).

To estimate the effect of travel restriction on spread, considering possible confounding effects of calendar date and the mediating effect of reported contacts of the index case, and to test the statistical significance of observed effects, a series of negative binomial regression models were fitted (Figure 5 and Supplementary Table 5). In the four weeks following the index case, fewer genomically-linked cases were reported when the index case was imported from a country with travel restrictions compared to cases from a non-travel restriction country (R.R.=0.17, 95% CI=0.05-0.52). When the number of contacts of the index case was included in the model, this rate ratio was attenuated slightly toward 1 (R.R.=0.25; 0.08-0.81) suggesting a limited mediating effect of the number of contacts of the index case. The effect of contacts was still seen, but the rate of subsequent cases (over four weeks) with the same genome was 4.0 (1.1-15.1) times higher for index cases with five or more reported contacts compared to those with none.

### Genomic identification of a large imported cluster

The Polecat Clustering tool (https://cog-uk.github.io/polecat) was used to analyse genomes in UK data on 14 September 2020. An outlier cluster was observed (Supplementary Figure 3). This cluster (UK1897) was associated with high diversity with a long stem length compared to samples from the UK, suggesting that this lineage evolved outside the UK. The geographic distribution of this lineage is demonstrated in Supplementary Figure 4, likely representing multiple importations across the UK (Supplementary Figure 4). This cluster contained the D614G mutation but no others associated with increased transmission. The root of the cluster was associated with a Swiss phylotype when linked to data in GISAID. During the course of the study period (04/08/2020 to 14/09/2020) there were 304 genomes. These were linked to 238 individuals, of whom 159 could be linked to a contact-tracing record. 143/159 had contact-tracing information indicating international travel or not. 72/143 (50.3%) individuals were linked to international travel and associated with ten, dispersed European countries (four individuals had travel to more than one European Country) and most commonly Croatia (35/72, 48.6%) (Supplementary Figure 5). A further 4 cases were identified as contacts of individuals who had reported travel to mainland Europe. There is a trend towards an increased proportion of cases that do not report travel over time, and possibly representing dispersion and onwards transmission locally of this lineage (Supplementary Figure 6).

### Lineage diversity of imported SARS-CoV-2 cases

The 827 imported genomes reflected 238 UK lineages (see Supplementary Materials), of which 214 were seen less than 5 times (142 singletons) and 24 were seen 5 or more times (Supplementary Table 6). The most commonly observed were UK5 (152 genomes, 18.4%) and UK1897 (73 genomes, 8.8%). There were 39 global lineages within the genomes. The most commonly observed lineages were B.1.1 (159 genomes, 19.2%) and B.1.177 (128 genomes, 15.5%) (Supplementary Tables 7 and 8). Further, potentially functionally important mutations were identified (Supplementary Table 9 and Supplementary Figure 8): D614G, 824/827 (99.6%) cases; N439K, 65/827 (7.86%) of cases; A222V, 131/827 (15.84%) of cases. ΔH69/V70 was identified in 53 cases associated with lineage B.1.258. We evaluated the introduction of A222V (B.1.177) over time, demonstrating a clear epidemiological link to Spain through contact tracing (Supplementary Figure 9). By the end of the study period, this variant was introduced from 16 separate countries indicating dispersion across Europe (Supplementary Figures 10). The mutations co-occur, with the proportion of cases represented by these combinations varying over time (Supplementary Figure 11).

## Discussion

We demonstrate, through the analysis of both contact-tracing data and the use of genomics, that travel restrictions (use of travel corridors) reduced the detected linked cases of SARS-COV-2. From 27/05/2020 to 13/09/2020, 85.9%% of importations were from European countries with three countries accounting for 51.2% of all imported cases. Along with travel restriction, age was a significant determinant of onwards contacts, and this effect was mitigated with closing travel corridors. After a period of national lockdown, systematic monitoring of imported genomes can identify sequences that are sufficiently unique and provide utility for monitoring of onwards transmission.

Whilst the study period covers nearly 5 months, the importations are concentrated after the implementation of travel corridors. The peaks for imports for each country occur at different times and with different epidemic curves. For the most common destination, barring Spain, imported cases appear to reduce after country-specific travel restrictions. Importations from Greece came at the end of August and continued into September, with the steepest of all curves. No travel restrictions were imposed on Greece during this time period and it was the source of greatest imported SARS-CoV-2 cases during this study period. This highlights the need for active surveillance of imported cases of SARS-CoV-2 for the introduction of travel corridors in a timely manner. London accounts for 15.4% of the population in England (19), but had 28.6% of the imports, possibly reflecting a younger age demographic. The overall R0 remained largely similar to other parts of the country during the study period potentially indicating imports are unlikely to have had a substantial impact on onward infection rates (20). Other explanations may include a small possible effect of higher seropositivity rates in London (17.5%, 27/4/2020, (21)) from the first wave of SARS-CoV-2 infections in England seen and a potential lower detection rate in London.

The number of onwards contacts are significantly reduced by the introduction of travel restrictions. Age is also a significant determinant of onwards contacts, with the 16-20 year old age-group representing the greatest number of travel-related cases and onwards contacts. This identifies an opportunity to direct public health awareness campaigns to younger travellers, with the intention to promote behaviours that will reduce the risk of SARS-CoV-2 acquisition and enhance compliance with quarantine on return to the UK.

The use of genomic sequencing, specifically after a period of national lockdown, allowed identification of a cohort of unique genomes that could be monitored for cluster growth. The cluster size for genomes that were related to a country without travel restrictions was significantly higher than those related to countries under travel restriction guidance. Further, when comparing the number of genomes in a cluster to the number of contacts that their respective cases reported, there was a trend towards a positive correlation suggesting self-isolation is effective. The total effect of travel restrictions was not explained by forward contacts alone and it is a possible that a reduction in the absolute number of individuals travelling to countries with travel restrictions also contributes to this.

The Polecat Clustering Tool highlighted a cluster that developed largely through travel to Croatia. Programmatic analysis of genomics data can therefore identify putative importation clusters. Integration with contact-tracing information was vital for the true picture of the sources of introduction and the subsequent spread, due to the SARS-CoV-2 sequencing bias observed globally. In this instance an introduced lineage was associated with wide-spread dispersal and onward transmission during a period when England had limited social distancing measures. The lineage, B.1.160, associated with this cluster is not associated with increased transmissibility but this study highlights a supplementary method for the detection and monitoring of expanding imported clusters and could prove particularly useful for the investigation of introduced variants of concern.

Our study is subject to multiple limitations. The COG-UK dataset has a limited sequencing coverage across England and cluster sizes detected will under-estimate absolute numbers (Supplementary Figure 12). The dates of country-specific travel restriction guidance was aligned with the date of travel-related case sampling, the earliest date reliably available. The effect of this should not however markedly affect results or conclusions; the period of travel restrictions are long and the effect size seen is large and therefore this discrepancy is unlikely to account for the significant difference observed. Further, most countries accounting for the imported SARS-CoV-2 cases went into a period of a travel restriction over the study period; by using a date later than the date of return from travel, we are more likely to over-represent contacts for countries under ‘travel-restriction’ guidance. Our study evaluates a period of time following a national lockdown and the associated reduced travel would likely exaggerate the diversity of genomes when compared with the COG-UK dataset. Outcomes such as travel and the number of contacts are self-reported by cases which will have inherent biases. Finally, there will be an artificial reduction in cases at the end of the study period when accounting for case incubation period, testing and report, with data provided 3 days after study close.

## Conclusions

We present an integrated epidemiological and genomic evaluation of the largest dataset of confirmed SARS-CoV-2 imported cases into the UK (or any other country) to our knowledge. We demonstrate the efficacy of closing ‘travel-corridors’ in reducing onward transmission of imported cases, and highlight the importance for targeted public health campaigns to reduce SARS-CoV-2 importations and onwards transmission. Our data demonstrates how routine genomic monitoring of travel-related cases could be used to refine travel restrictions and the genomic diversity of the SARS-CoV-2 import cases.

## Supporting information

COVID-19 Genomics UK (COG-UK) Consortium authorship

## Data Availability

Assembled/consensus genomes are available from GISAID (22) subject to minimum quality control criteria. Raw reads are available from European Nucleotide Archive (ENA) (23). All genomes, phylogenetic trees, basic metadata are available from the COG-UK consortium website (https://www.cogconsortium.uk/data). For confidentiality reasons, extended metadata (24) is not publicly available, however some may be available upon request from Public Health England.

## Author contributions

All authors read the manuscript and consented to its publication.

DA led the study.

DA, AJP, GS wrote the first draft of the manuscript.

All authors contributed to revision of the first draft of the manuscript.

DA, US, AJP, RM, NE undertook data analysis.

GMS provided statistical guidance and analysis.

DA, SP, NG, EG contributed to data curation.

GJH, CC, CT, SL, AH provided oversight over data acquisition and data definitions.

TLV wrote scripts to perform analysis.

NMT undertook the literature search.

DA, US, RM, EV, MC, EMH contributed to study design

MC, EMH, SJP conceived the study and provided overall leadership.

DA, MC and SJP provided clinical oversight.

## Ethics

This study was conducted as part of surveillance for COVID-19 infections under the auspices of Section 251 of the NHS Act 2006 and/or Regulation 3 of The Health Service (Control of Patient Information) Regulations 2002. They therefore did not require individual patient consent or ethical approval. Public Health England affiliated authors had access to identifiable patient data. Other authors only had access to anonymised or summerised data. The COG-UK study protocol was approved by the Public Health England Research Ethics Governance Group (reference: R&D NR0195).

## Acknowledgements

We thank members of the COVID-19 Genomics Consortium UK and Test and Trace contact tracers or their contributions to generating the data used in this study. We thank Sarah Mitchell from the Department of Plant Sciences, University of Cambridge, for direction on statistical analysis.

