## Supplementary Tables, Figures and Methods for "An integrated analysis of contact tracing and genomics to assess the efficacy of travel restrictions on SARS-CoV-2 introduction and transmission in England from June to September, 2020"

| Study | Month (2020) | Country | Imported from | No. imports | Genomes |
|---|---|---|---|---|---|
| (25) | March | China | US (2), Germany (1) | 3 | 7 |
| (26) | January | Germany | China | 1 | |
| *(27)* | | Israel | Japan (1), Italy (1) | 2 | |
| (28) | | Brazil | Italy (4) | | 6 |
| (29) | March | Brazil | Spain | 1 | |
| (30) | Before May | China | South America (2), North America (15), Europe (101), Other Asian countries (3) | | 102 |
| (31) | February | Mexico | Italy | 1 | |
| *(32)* | February - March | South Africa | Europe | 13 | 27 |
| (33) | January | Italy | China | 2 | |
| (34) | Before June | Spain | | > 34 | |
| (35) | January - March | Taiwan | China, Germany, UK, Turkey, Iran, Middle East, Europe | | 20 |
| *(36)* | March | China | Spain (2), France (1), Cambodia (1), Sri Lanka (1), US (3) | 8 | |
| *(37)* | April | Mali | | | 21 |
| (38) | | India | China, South Asia, Middle East, Italy, Spain, UK, France and USA | | 104 |
| (39) | April | China | US | 1 | |
| (40) | January - March | China | 19 different countries | 102 | 53 new + 177 publicly available sequences |
| (41) | January | Cambodia | | 1 | |
| (42) | March | Ecuador | Netherlands | 1 | 4 |
| (43) | <May | Thailand | China | 1 | 40 |
| (44) | January - March | Australia | Asia, Western Europe and North America | | |
| (45) | | Australia | Asia, Europe, North America | 193 | 76 clusters, 34 only international travel, 34 mixed local/international |
| (46) | March | Japan | Egypt | 10 | 26 |
| (47) | February - March | Switzerland | Italy (2), France (1), Austria (refers to a previous study) | | 486 |
| (Tapfumani et al 2020) | February - March | Zimbabwe | UK, US, South Africa, Dubai | | 97 |

**Supplementary Table 1: Studies using genomics to as part of epidemiological investigations of importations of SARS-CoV-2.**

| Ethnic group | Cases | | Census |
|---|---|---|---|
| **White** | **No.** | **%** | **%** |
| English/Welsh/Scottish/Northern Irish/British | 2509 | 66.8% | 80.5% |
| Irish | 35 | 0.9% | 0.9% |
| Gypsy or Irish Traveller | 0 | 0.0% | 0.1% |
| Any other White background | 583 | 15.5% | 4.4% |
| **Mixed/Multiple ethnic groups** | | | |
| White and Black Caribbean | 42 | 1.1% | 0.8% |
| White and Black African | 25 | 0.7% | 0.3% |
| White and Asian | 49 | 1.3% | 0.6% |
| Any other Mixed/Multiple ethnic background | 44 | 1.2% | 0.5% |
| **Asian/Asian British** | | | |
| Indian | 103 | 2.7% | 2.5% |
| Pakistani | 79 | 2.1% | 2.0% |
| Bangladeshi | 27 | 0.7% | 0.8% |
| Chinese | 6 | 0.2% | 0.7% |
| Any other Asian background | 62 | 1.7% | 1.5% |
| **Black/ African/Caribbean/Black British** | | | |
| African | 58 | 1.5% | 1.8% |
| Caribbean | 12 | 0.3% | 1.1% |
| Any other Black/African/Caribbean background | 12 | 0.3% | 0.5% |
| **Other ethnic group** | | | |
| Arab | 0 | 0.0% | 0.4% |
| Any other ethnic group | 110 | 2.9% | 0.6% |
| **Other** | | | |
| Prefer not to say | 65 | | |
| Unknown | 386 | | |

**Supplementary Table 2: Self-identified ethnicity of cases (UK Government Statistical Service ethnic groups). The 2011 census data for England and Wales was used.**

| Demographic | Cases | | Contacts | | Cases with Genomes that passed QC | |
|---|---|---|---|---|---|---|
| **Sex** | | | | | | |
| Male | 2193 | 56.2% | 6088 | 49.7% | 394 | 47.8% |
| Female | 1933 | 46.8% | 6160 | 50.3% | 414 | 50.2% |
| Unknown | 81 | | 6607 | | 19 | |
| **Age** | | | | | | |
| 0-5 | 51 | 1.2% | 303 | 2.5% | 9 | 1.1% |
| 6-10 | 45 | 1.1% | 361 | 2.9% | 12 | 1.5% |

| | | | | | | |
|---|---|---|---|---|---|---|
| 11-15 | 75 | 1.8% | 467 | 3.8% | 14 | 1.7% |
| 16-20 | 1086 | 25.8% | 2274 | 18.5% | 228 | 27.6% |
| 21-25 | 843 | 20.0% | 1866 | 15.2% | 165 | 20.0% |
| 26-30 | 685 | 16.3% | 1350 | 11.0% | 135 | 16.3% |
| 31-35 | 413 | 9.8% | 849 | 6.9% | 88 | 10.6% |
| 36-40 | 278 | 6.6% | 721 | 5.9% | 46 | 5.6% |
| 41-45 | 185 | 4.4% | 691 | 5.6% | 35 | 4.2% |
| 46-50 | 169 | 4.0% | 984 | 8.0% | 34 | 4.1% |
| 51-55 | 130 | 3.1% | 1168 | 9.5% | 22 | 2.7% |
| 56-60 | 121 | 2.9% | 692 | 5.6% | 23 | 2.8% |
| 61-65 | 57 | 1.4% | 264 | 2.1% | 7 | 0.8% |
| 66-70 | 30 | 0.7% | 141 | 1.1% | 4 | 0.5% |
| 71-75 | 20 | 0.5% | 101 | 0.8% | 4 | 0.5% |
| 76-80 | 7 | 0.2% | 35 | 0.3% | 0 | 0.0% |
| 81-85 | 7 | 0.2% | 30 | 0.2% | 1 | 0.1% |
| 86-90 | 3 | 0.1% | 7 | 0.1% | 0 | 0.0% |
| 91-95 | 0 | 0.0% | 4 | 0.0% | 0 | 0.0% |
| Unknown | 2 | | 6547 | | 0 | |
| **Ethnic group** | | | | | | |
| **White** | | | | | | |
| English/Welsh/Scottish/Northern Irish/British | 2509 | 66.8% | 8370 | 73.8% | 499 | 68.4% |
| Irish | 35 | 0.9% | 134 | 1.2% | 4 | 0.5% |
| Gypsy or Irish Traveller | 0 | 0.0% | 0 | 0.0% | 0 | 0.0% |
| Any other White background | 583 | 15.5% | 1261 | 11.1% | 120 | 16.4% |
| **Mixed/Multiple ethnic groups** | | | | | | |
| White and Black Caribbean | 42 | 1.1% | 88 | 0.8% | 4 | 0.5% |
| White and Black African | 25 | 0.7% | 60 | 0.5% | 8 | 1.1% |
| White and Asian | 49 | 1.3% | 112 | 1.0% | 9 | 1.2% |
| Any other Mixed/Multiple ethnic background | 44 | 1.2% | 124 | 1.1% | 9 | 1.2% |
| **Asian/Asian British** | | | | | | |
| Indian | 103 | 2.7% | 318 | 2.8% | 13 | 1.8% |
| Pakistani | 79 | 2.1% | 183 | 1.6% | 4 | 0.5% |
| Bangladeshi | 27 | 0.7% | 50 | 0.4% | 3 | 0.4% |
| Chinese | 6 | 0.2% | 33 | 0.3% | 1 | 0.1% |
| Any other Asian background | 62 | 1.7% | 197 | 1.7% | 13 | 1.8% |
| **Black/ African/Caribbean/Black British** | | | | | | |
| African | 58 | 1.5% | 148 | 1.3% | 12 | 1.6% |
| Caribbean | 12 | 0.3% | 35 | 0.3% | 3 | 0.4% |
| Any other Black/African/Caribbean background | 12 | 0.3% | 25 | 0.2% | 2 | 0.3% |
| **Other ethnic group** | | | | | | |

| | | | | | | |
|---|---|---|---|---|---|---|
| Arab | 0 | 0.0 | 0 | 0.0% | 0 | 0.0% |
| Any other ethnic group | 110 | 2.9% | 195 | 1.7% | 26 | 3.6% |
| **Other** | | | | | | |
| Prefer not to say | 65 | | 139 | | 85 | |
| Unknown | 386 | | 7382 | | 12 | |
| **Region** | | | | | | |
| London | 1205 | 28.6% | 4681 | 24.9% | 298 | 36.3% |
| South East | 623 | 14.8% | 3201 | 17.0% | 175 | 21.3% |
| North West | 584 | 13.9% | 2503 | 13.3% | 70 | 8.5% |
| East of England | 395 | 9.4% | 1958 | 10.4% | 94 | 11.4% |
| South West | 328 | 7.8% | 1755 | 9.3% | 74 | 9.0% |
| Yorkshire and Humber | 327 | 7.8% | 1488 | 7.9% | 21 | 2.6% |
| West Midlands | 299 | 7.1% | 1410 | 7.5% | 20 | 2.4% |
| East Midlands | 251 | 6.0% | 1157 | 6.2% | 29 | 3.5% |
| North East | 161 | 3.8% | 646 | 3.4% | 40 | 4.9% |
| Not stated | 34 | 0.8% | 56 | | 6 | |

**Supplementary Table 3: Demographics of cases, contacts, and cases with genomes that pass quality control available**

| | **Effect of travel restriction (ratio of mean contacts)** | **Adjusted mean contacts** | |
|---|---|---|---|
| | | **With travel restriction** | **Without travel restriction** |
| **Overall** | 0.60 (0.37-0.95) | 3.50 (3.04-4.02) | 5.85 (3.67-9.34) |
| **By age-group** | | | |
| 0-15 | 0.73 (0.39-1.34) | 4.3 (3.3-5.6) | 5.9 (3.3-10.3) |
| 16-20 | 0.52 (0.32-0.85) | 4.7 (3.9-5.7) | 9.0 (5.6-14.5) |
| 21-25 | 0.54 (0.33-0.88) | 3.5 (2.9-4.2) | 6.5 (4.0-10.5) |
| 26-30 | 0.49 (0.30-0.80) | 2.4 (2.0-2.9) | 4.8 (3.0-7.8) |
| 31-40 | 0.58 (0.36-0.94) | 3.0 (2.5-3.6) | 5.2 (3.2-8.3) |
| 41 and older | 0.78 (0.48-1.29) | 3.7 (3.1-4.3) | 4.7 (2.9-7.6) |
| **By calendar date** | | | |
| May/June | | 5.9 (4.3-8.2) | Insufficient data |
| July | 0.72 (0.51-1.03) | 5.1 (4.0-6.4) | 7.0 (5.2-9.5) |
| August 1-14 | 0.42 (0.33-0.53) | 2.5 (2.1-3.1) | 6.1 (5.0-7.4) |
| August 15-31 | 0.60 (0.52-0.69) | 2.5 (2.1-2.8) | 4.1 (3.5-4.8) |
| September | 0.78 (0.65-0.93) | 2.8 (2.4-3.2) | 3.6 (3.0-4.2) |

**Supplementary Table 4: The effect of travel restriction on reported contacts per imported case, and the estimated marginal mean number of reported cases when imported from a country with or without a travel restriction in place.** Figures are reported for the overall dataset, and then stratified by age-group and calendar date of positive test. All estimates are provided with 95% confidence intervals

| | Effect of travel restriction on subsequent cases with matching genome (rate ratio with 95% confidence interval) | |
|---|---|---|
| | **2 weeks after index case** | **4 weeks after index case** |
| **Model without contacts** | | |
| Quarantine | 0.18 (0.06-0.55) | 0.17 (0.05-0.52) |
| **Model including contacts** | | |
| Quarantine | 0.27 (0.09-0.77) | 0.25 (0.08-0.81) |
| 1-4 contacts (vs none) | 1.03 (0.30-3.58) | 1.26 (0.34-4.75) |
| 5 or more contacts (vs none) | 4.14 (1.14-15.02) | 4.01 (1.06-15.1) |

**Supplementary Table 5: Effect of travel restriction on number of genomes detected.** Effect of travel restriction on number of genomes detected (measured by rate ratio with 95% confidence interval). All estimates are adjusted for calendar date.

| Lineage | No. Samples | Percentage |
|---|---|---|
| UK5 | 152 | 18.4% |
| UK1897 | 73 | 8.8% |
| UK461 | 66 | 8.0% |
| UK2229 | 28 | 3.4% |
| UK1249 | 22 | 2.7% |
| UK649 | 22 | 2.7% |
| UK1506 | 22 | 2.7% |
| UK1031 | 12 | 1.5% |
| UK1205 | 10 | 1.2% |
| UK2347 | 10 | 1.2% |
| UK761 | 10 | 1.2% |
| UK1780 | 8 | 1.0% |
| UK1791 | 8 | 1.0% |
| UK1569 | 7 | 0.8% |
| UK831 | 7 | 0.8% |
| UK669 | 7 | 0.8% |
| UK1018 | 6 | 0.7% |
| UK1219 | 6 | 0.7% |
| UK2683 | 6 | 0.7% |
| UK2726 | 6 | 0.7% |
| UK778 | 5 | 0.6% |
| UK1535 | 5 | 0.6% |
| UK1581 | 5 | 0.6% |
| UK2268 | 5 | 0.6% |
| 214 lineages with <5 cases | 319 | 38.6% |

**Supplementary Table 6: The number of samples with each UK lineage**

| Lineage | No. Samples | Percentage |
|---|---|---|
| B.1.1 | 159 | 19.2% |
| B.1.177 | 128 | 15.5% |
| D.1 | 87 | 10.5% |
| B.1.160 | 75 | 9.1% |
| B.1.5 | 72 | 8.7% |
| B.1 | 72 | 8.7% |
| B.1.1.1 | 55 | 6.7% |
| B.1.1.37 | 55 | 6.7% |
| B.1.78 | 36 | 4.4% |
| B.1.1.70 | 17 | 2.1% |
| B.1.36 | 14 | 1.7% |
| B.1.5.12 | 6 | 0.7% |
| B.1.36.1 | 6 | 0.7% |
| B.1.1.34 | 5 | 0.6% |
| 25 lineages with <5 cases | 40 | 4.8% |

**Supplementary Table 7: The number of travel-related samples with each Global lineage**

| Lineage | Number | Percentage |
|---|---|---|
| B.1.1 | 7673 | 37.2 |
| B.1.177 | 1862 | 9.0 |
| B.1 | 1547 | 7.5 |
| B.1.5 | 1299 | 6.3 |
| B.1.1.37 | 1173 | 5.7 |
| B.1.1.35 | 996 | 4.8 |
| B.1.1.1 | 805 | 3.9 |
| D.1 | 647 | 3.1 |
| B.1.160 | 405 | 2.0 |
| B.1.36.1 | 362 | 1.8 |
| B.1.1.4 | 335 | 1.6 |
| B | 321 | 1.6 |
| B.1.1.51 | 319 | 1.5 |
| B.1.1.30 | 233 | 1.1 |
| B.1.36 | 232 | 1.1 |
| B.1.1.15 | 189 | 0.9 |
| B.1.78 | 181 | 0.9 |
| B.1.1.55 | 145 | 0.7 |
| C.3 | 125 | 0.6 |
| B.1.1.70 | 112 | 0.5 |

**Supplementary Table 8: The number of samples with each Global lineage from the COG-UK dataset during the study period.** This table includes the 'top 20' lineages sequenced during the study period.

| Mutation | Cases with Mutant Variant | | Wild Type Variant | | Inconclusive | |
|---|---|---|---|---|---|---|
| D614G | 824 | 99.64% | 3 | 0.36% | 0 | 0.00% |
| P323L* | 4 | 0.48% | 815 | 98.55% | 7 | 0.85% |
| N439K | 65 | 7.86% | 758 | 91.66% | 4 | 0.48% |
| A222V | 131 | 15.84% | 694 | 83.92% | 2 | 0.24% |
| Y453F | 0 | 0.00% | 826 | 99.88% | 1 | 0.12% |
| Total Cases | 1114 | | | | | |

**Supplementary Table 9: Mutant variants identified in the travel-related cases during the study period. *F mutation found in 1 case**

**Supplementary Figures**

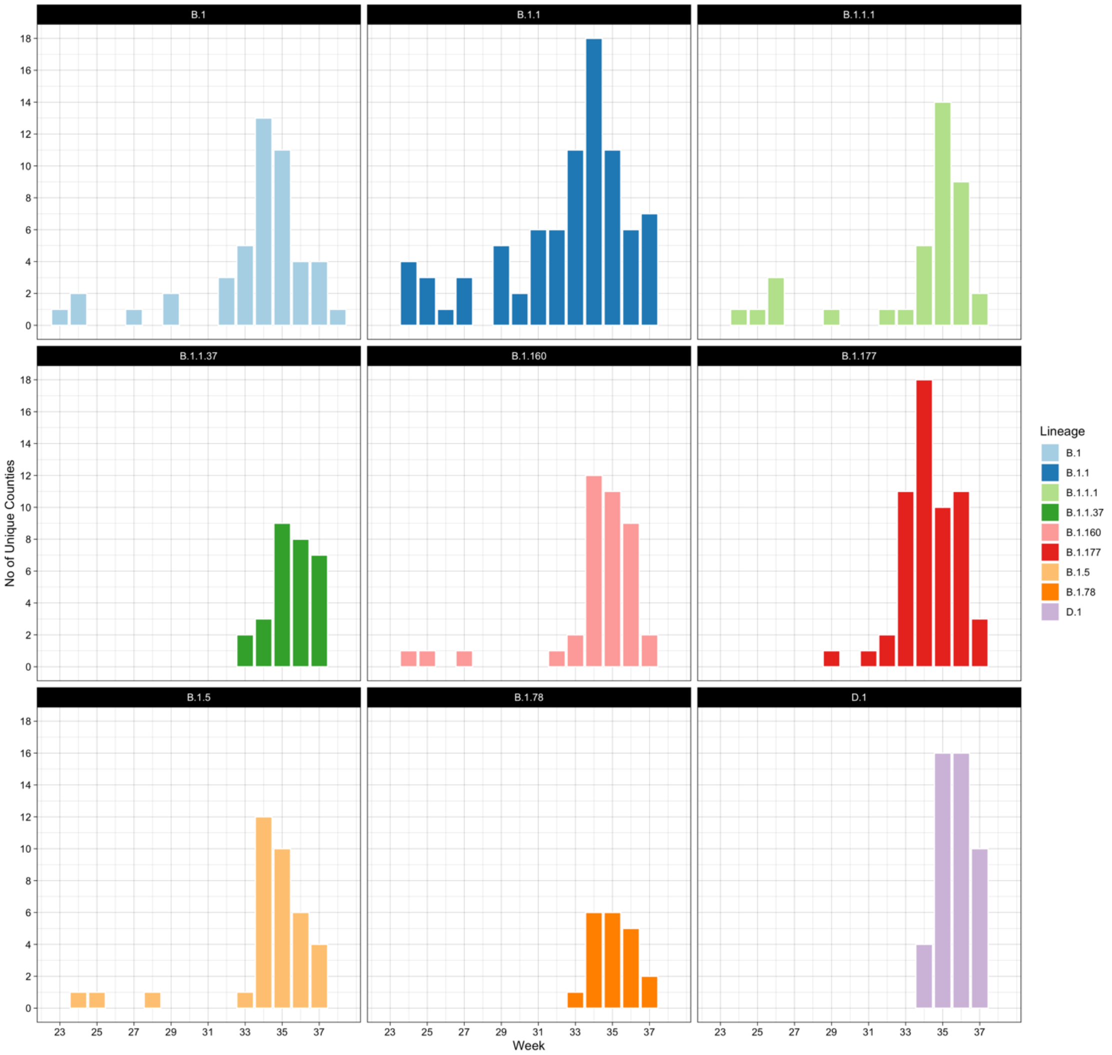


**Supplementary Figure 1: The dispersion of importations of different lineages throughout England per week.** This represents the top 9 global lineages versus the number of unique counties the lineage is found in, using the county provided by the case. The counties are the lieutenancies or ceremonial counties of which there are 48.

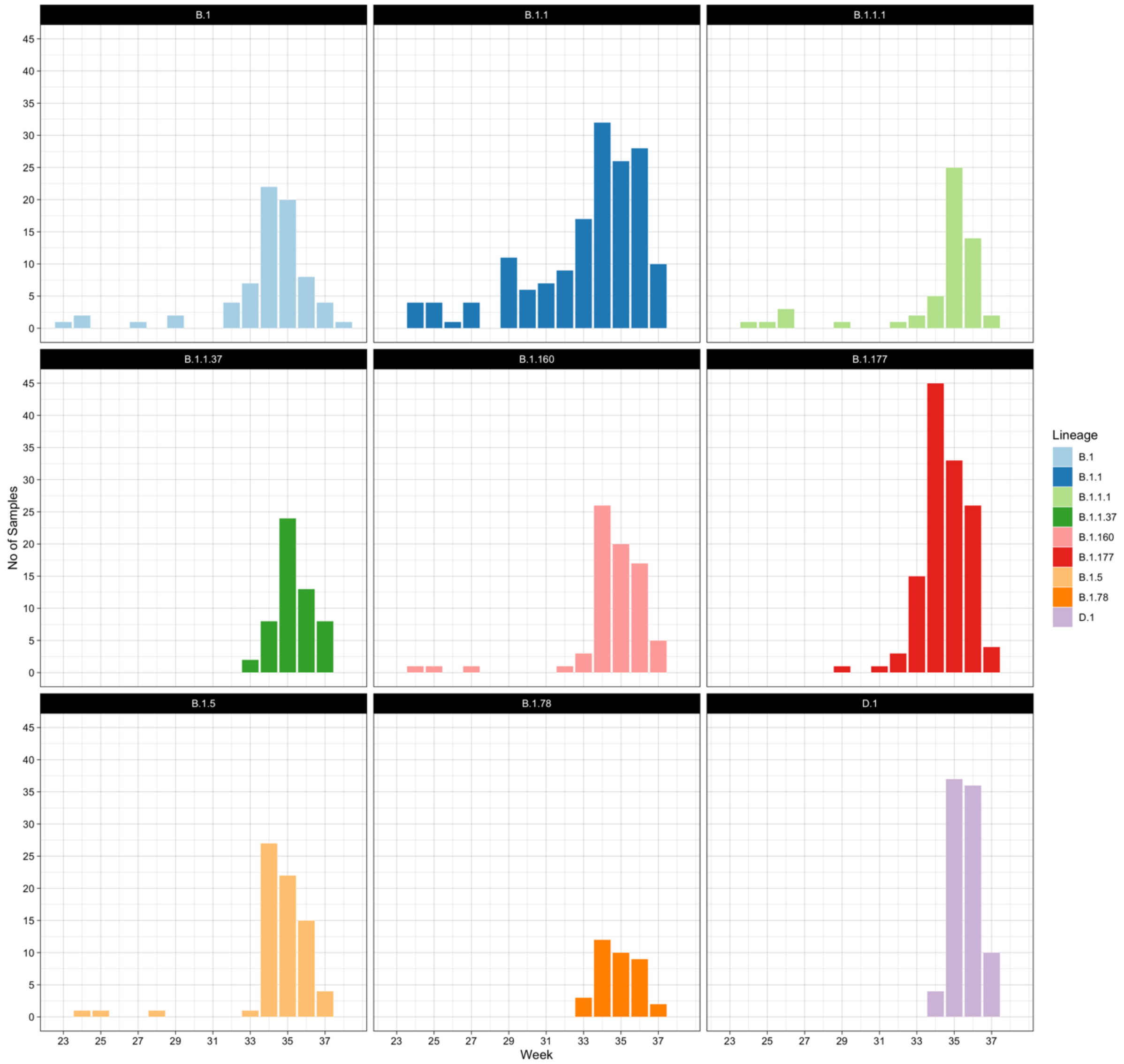


**Supplementary Figure 2: The number of importations of each global lineage per week of 2020.** This Figure represents the Top 9 global lineages.

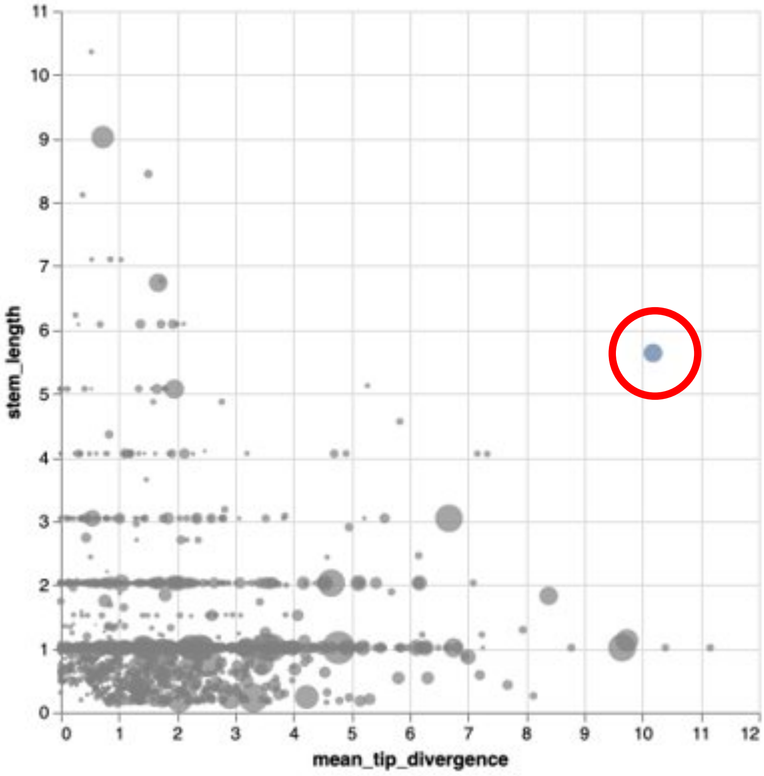


**Supplementary Figure 3: Polecat cluster analysis with the likely travel-related cluster highlighted.**

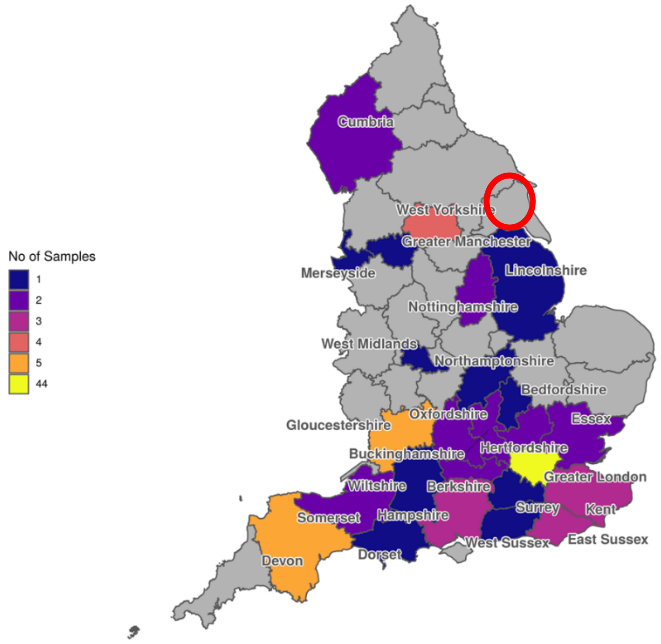


**Supplementary Figure 4: No. of genomes of importations or their contacts of lineage UK1897 per county in England.**

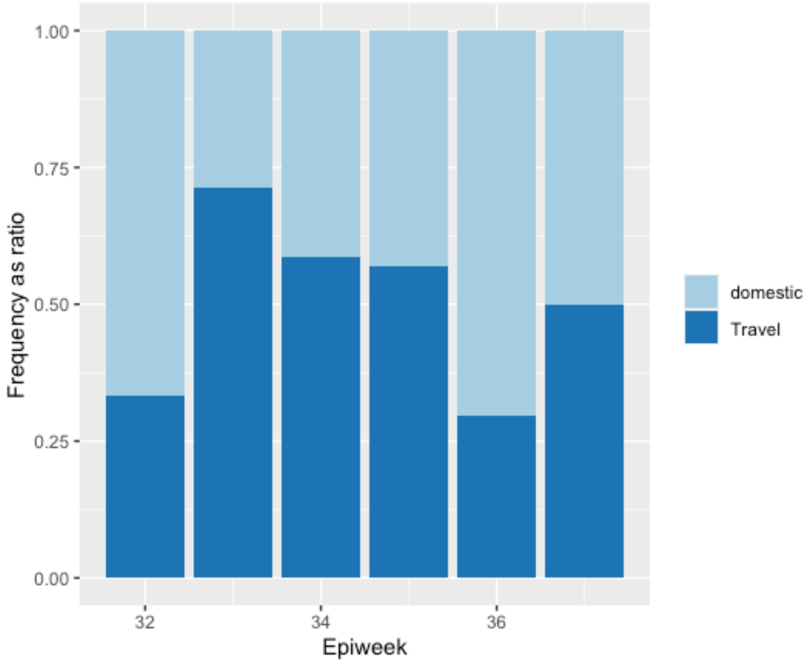


**Supplementary Figure 5: Frequency of individuals identifying a travel or domestic source of SARS-CoV-2 acquisition within the suspected travel-related cluster of genomes highlighted by the Polecat tool, represented by epiweek**

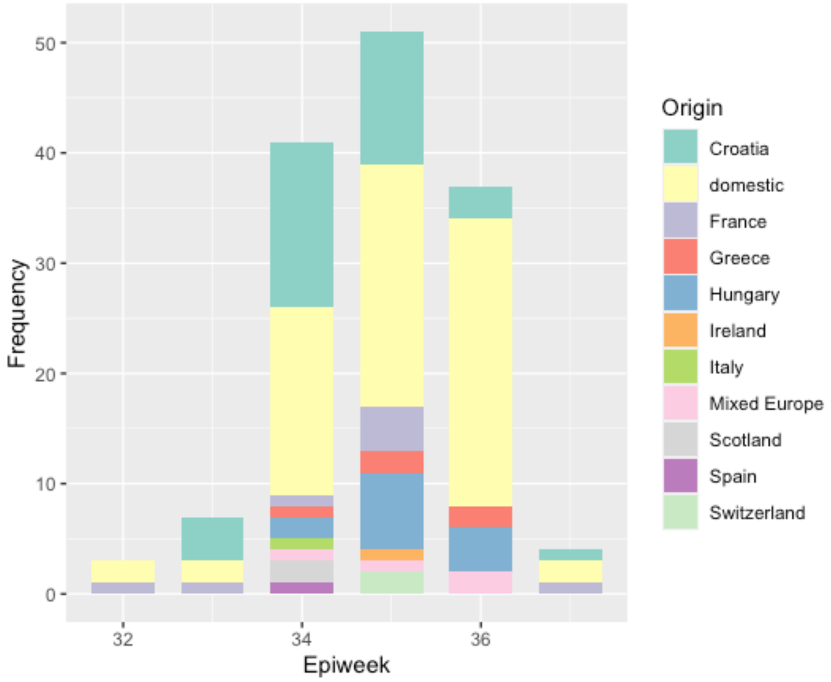


**Supplementary Figure 6: Source country of SARS-CoV-2 acquisition for individuals identified within a suspected travel-related cluster highlighted by the Polecat tool, represented by epiweek**

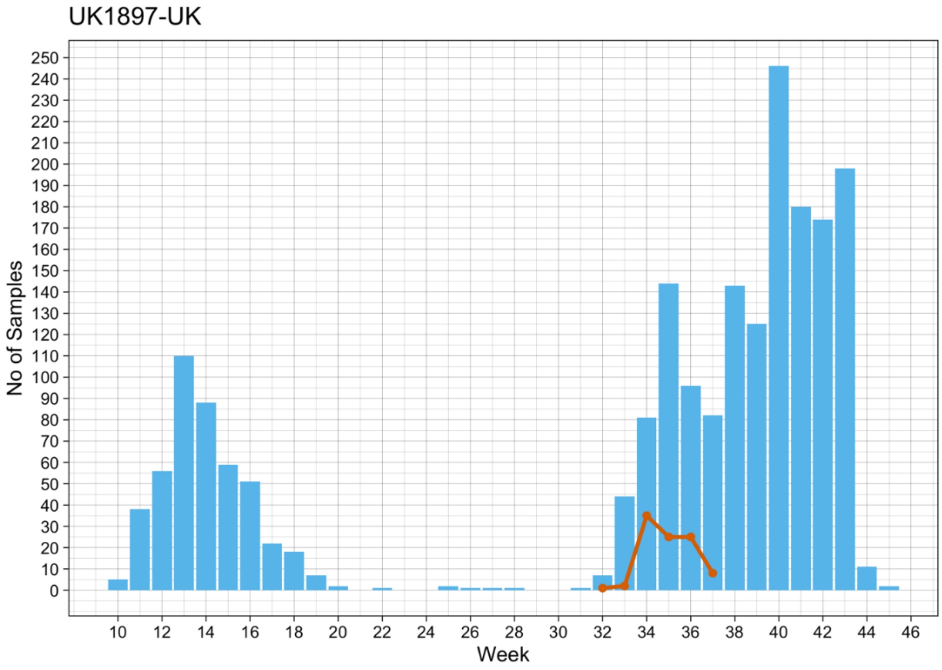


**Supplementary Figure 7: UK1897 SARS-CoV-2 lineage in the United Kingdom by epiweek.** The line (orange) is the number of genomes which are confirmed importations from the lineage UK1897 per week of 2020. The blue bars indicate the number of genomes of this lineage seen per week anywhere in the UK (including the importations).

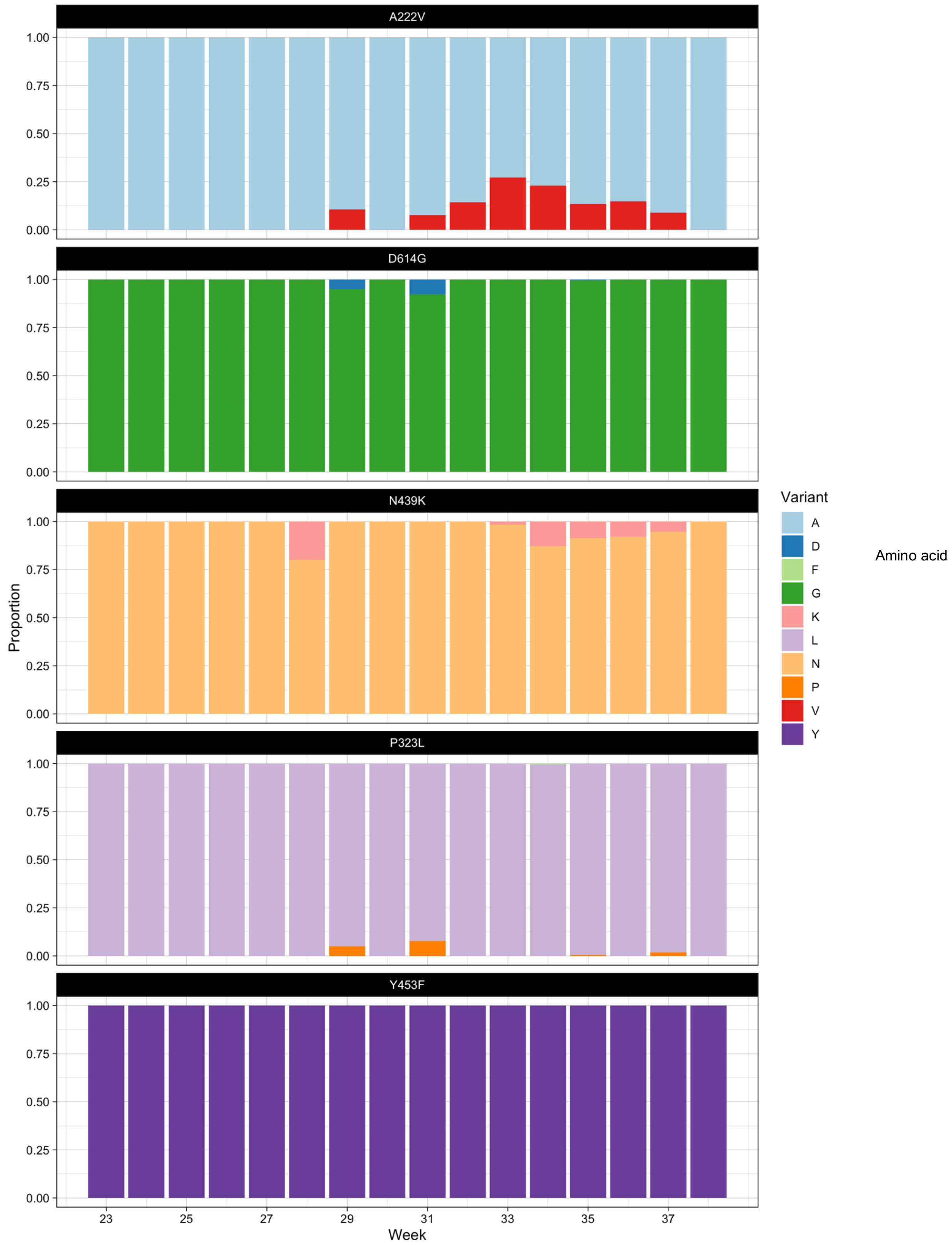


**Supplementary Figure 8: The percentage of each major mutation observed per week in the imported genomes.** Letters in the amino acid substitution nomenclature correspond to: A, alanine; D, aspartic acid; F, phenylalanine; G, glycine; K, lysine; L, leucine; N, asparagine; P, proline; V, valine; Y, tyrosine. The mutations are named as following: the letter preceding number (the amino acid site of substitution) represents the wild-type amino acid, the letter following the number is the observed amino acid in the sample ('a mutation', if different from the wild-type). The figure legend represents the observed amino acid at the site of interest, e.g. 'A' in the panel representing the A222V mutation shows cases observing alanine at site 222.

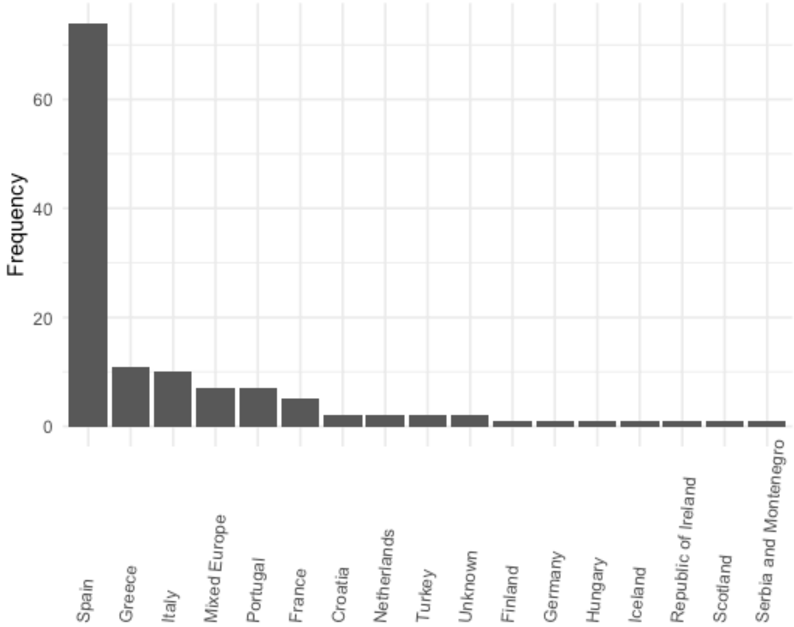


**Supplementary Figure 9: Destination country of travel-related SARS-CoV-2 with the A222V variant identified, during the study period**

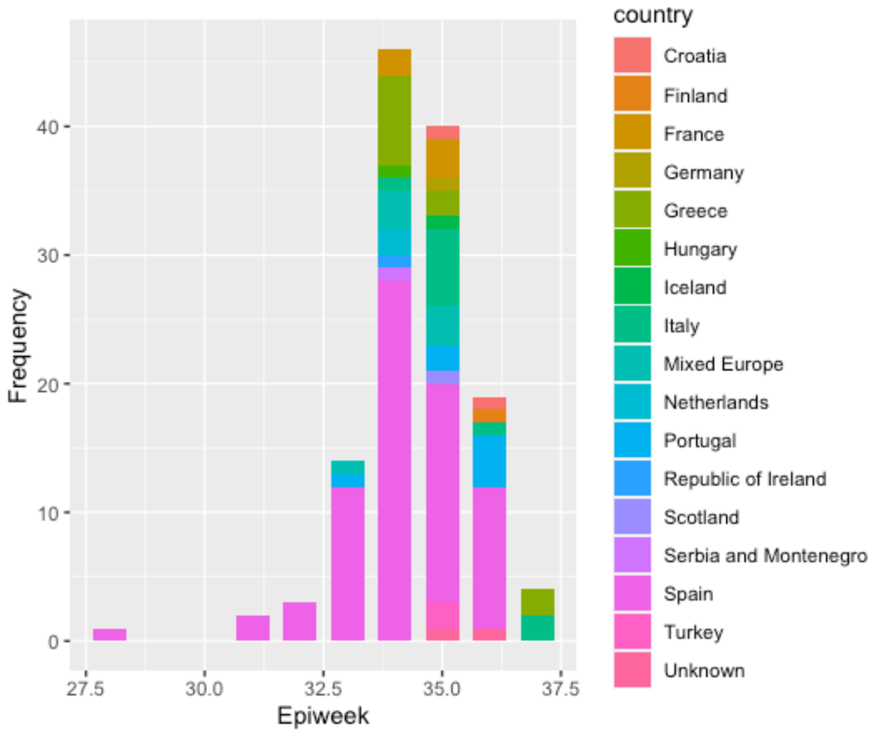


**Supplementary Figure 10: Reported country of travel for cases with the A222V variant of SARS-CoV-2, imported over time**

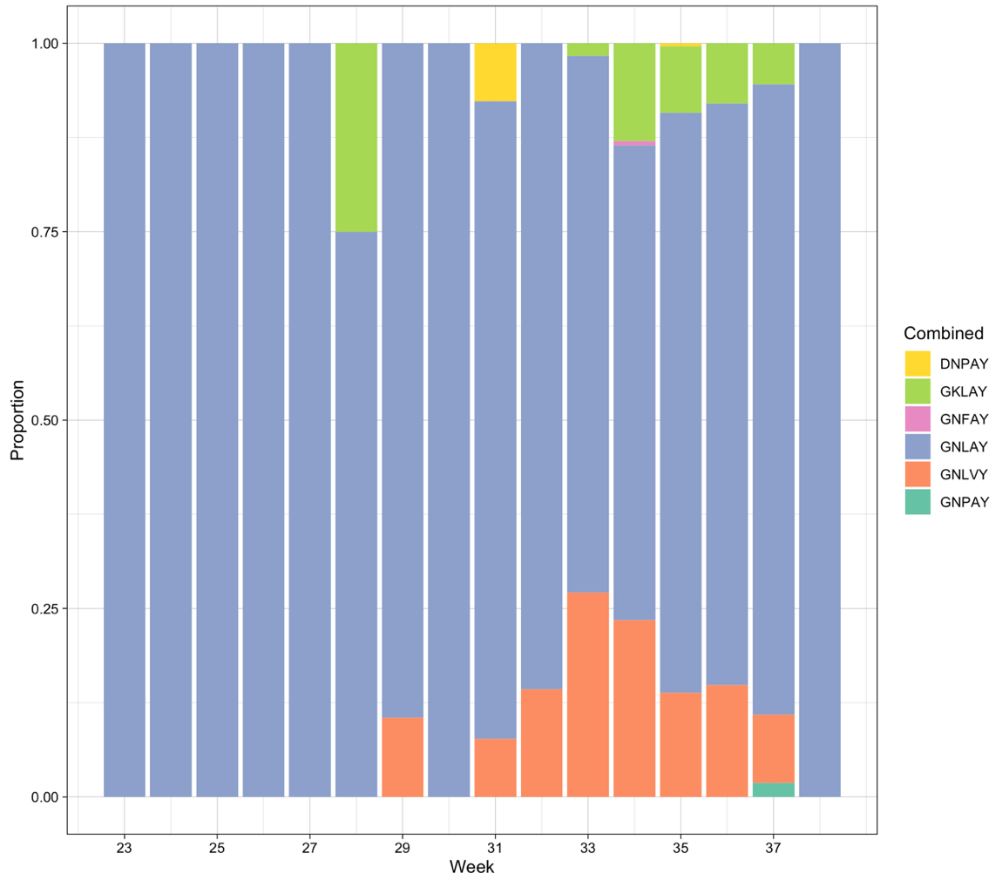


**Supplementary Figure 11: Combination of SARS-CoV-2 mutations seen in imported SARS-CoV-2 genomes by epiweek.** The combinations of co-occurring variants, where the variants are in the order: 1) D614G, 2) N439K,3) P323L, 4) A222V and 5) Y453F.

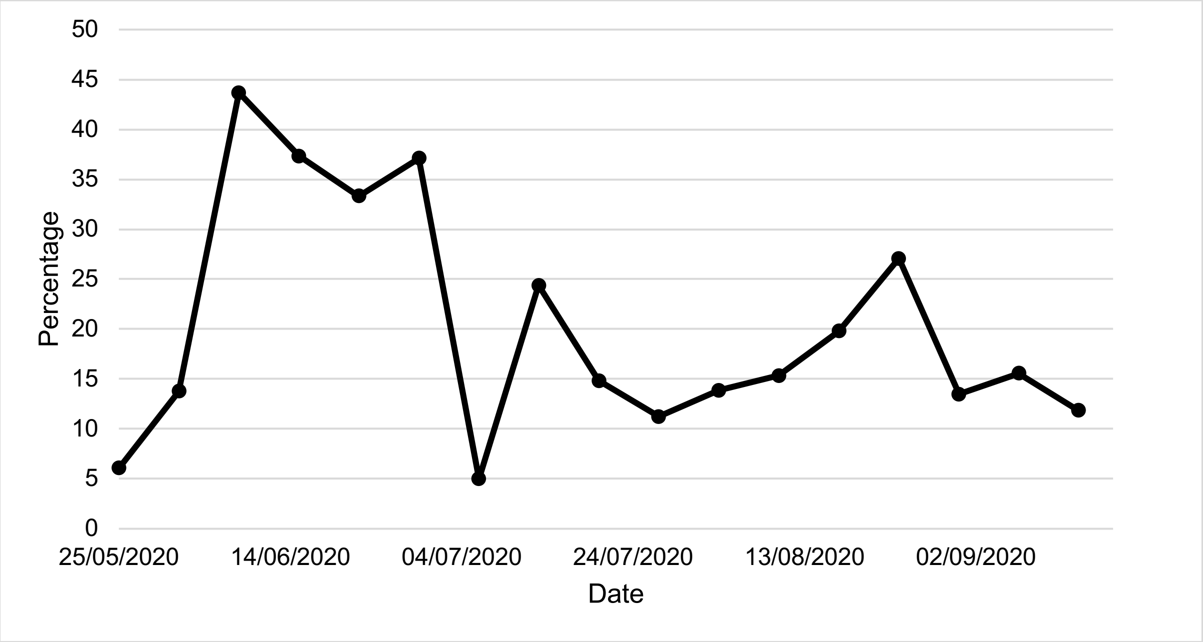


**Supplementary Figure 12: Percentage of known SARS-CoV-2 cases sequenced in England from 25/05/2020 to 14/09/2020**

### Supplementary Methods and Definitions

**Travel guidance**

During the time period of the study all non-essential travel outside of the UK was advised against. Varying restrictions were applied to travellers returning from different countries or regions of countries, changing over the course of the study period. Travellers were required to quarantine for 2 weeks if they had visited a restricted region in the previous 2 weeks. There were exceptions for particular classes of individuals such as freight drivers and flight crews. Designated regions were exempt from the quarantine requirements, and commonly referred to as open 'travel-corridors'. These restrictions changed over time for different regions (https://www.gov.uk/guidance/travel-advice-novel-coronavirus).

**Contact tracing and case identification**

Contact-tracing data was obtained from T&T. Case data gathered from testing laboratories is enriched with data provided by NHS Spine, prior to arrival at the contact tracing advise service system. All cases and contacts had a field for demographic data, but this was not always reported (Table 2 and Supplementary Table 3). 'Highly probable' travel-related cases were defined as individuals who reported international travel as an activity in the two days before symptom onset/testing. On 12/08/2020 the additional facility to report international travel in the seven days prior to symptom onset/testing became available, and also included in this study and defined as 'probable' travel-related cases.

Cases are asked to provide details of all contacts for activities in the 2 days prior to onset/testing up to completing the system which were gathered. If any contacts become cases they would then also be included in T&T as a case separately but if they did not report direct travel themselves, then they would not meet the definition for a travel-associated case.

Cases identified reporting travel in 7-day period prior to symptom onset or positive test:
Test and Trace data included destination city, and a free-text search was run with a custom python script to convert city to associated country of destination. All fields were manually cross-checked and any errors corrected (142 corrections). A further 103 countries were manually inputted due to spelling errors in the free-text Test and Trace data provided and 1 country by searching flight numbers provided by the case when country or city not available. 22 cases reporting travel-related activities did not have an associated destination clearly identified.

Cases identified reporting travel in 2-day period prior to symptom onset or positive test:
A free-text country and city search with a custom python script on travel-related T&T data was used to identify destination country. This yielded 1898 destination countries, and a further 1182 by city search. All fields were manually cross-checked and errors corrected (210 corrected). 542 case-country associations were manually entered where spelling mistakes were present in the free-text entries, including 98 entered by flight number searches where this was the only available data.

**Lineages**

Global and UK Lineages (14) were assigned to each genome using Pangolin (https://github.com/cov-lineages/pangolin) with analysis performed on COVID-CLIMB (13). Global lineages, reflecting genomically distinct identifiable importations into a new region, are denoted with a letter followed by a hierarchy of up to 4 numbers such as B.1.2.3, providing for a stable and consistent naming of clusters. These lineages are manually curated and assigned. UK lineages represent the subsequent regional and local spread within the UK, taking the form UK1234, providing an identifier for a cluster for a given phylogeny. These identifiers are assigned programmatically are unstable. Labelled phylogenetic trees were created using CIVET tool (version 2.0) (https://github.com/cog-uk/civet).

**Identification of extinct and unique genomes**

The 827 high-quality travel-related genomes were compared to the COG-UK dataset on 16/10/2020. Genomes were only compared to other genomes with the same UK lineage assigned by COG-UK, since we assume that no relatedness relevant to transmission exists between genomes of different UK lineages. A unique genome in the community was deemed to be one that was known to be from a travel-related case and either: (1) A UK lineage that had not been sampled in the previous 4 weeks in the UK, (2) >3 SNPs distance to the closest relative in the COG-UK dataset.
Within the same UK lineage we identified those genomes sampled within 4 weeks prior to the genome of interest. We determined the minimum SNP distance between the sequence of interest and these genomes. This identified 207/827 genomes with a minimum SNP distance of >3 SNPs to its closest relative in the COG-UK dataset. These constitute genomes for which no close relative was sampled in the UK at the time of importation. The analysis was then repeated on 05/12/2020 on these 207 'unique genomes' to account for delays in genomes uploaded to MRC CLIMB. 195/207 were included in this analysis, with 12/207 genomes excluded due to the large UK phylotypes they belonged to and the subsequent computational requirements. At this time a further 8 genomes were determined to have a close relative sampled in the UK in the 4 weeks before importation. This wasn't detected earlier, because their close relative was uploaded with a significant delay.
The remaining 186 genomes were 'Unique' genomes were compared to sequences that were generated in the COG-UK dataset within 2 and 4 weeks after their sampling date, to identify samples with the same UK lineage and within 2 SNPs. These would represent onward transmission or further introductions of similar genomes. The analysis was run with an in-house custom Python script developed by US and RM.
