## Supplementary material for "An integrated analysis of contact tracing and genomics to assess the efficacy of travel restrictions on SARS-CoV-2 introduction and transmission in England from June to September, 2020": COVID-19 Genomics UK (COG-UK) Consortium authorship

**Funding acquisition, Leadership and supervision, Metadata curation, Project administration, Samples and logistics, Sequencing and analysis, Software and analysis tools, and Visualisation:**
Dr Samuel C Robson [13].

**Funding acquisition, Leadership and supervision, Metadata curation, Project administration, Samples and logistics, Sequencing and analysis, and Software and analysis tools:**
Prof Nicholas J Loman [41], Dr Thomas R Connor [10, 69].

**Leadership and supervision, Metadata curation, Project administration, Samples and logistics, Sequencing and analysis, Software and analysis tools, and Visualisation:**
Dr Tanya Golubchik [5].

**Funding acquisition, Metadata curation, Samples and logistics, Sequencing and analysis, Software and analysis tools, and Visualisation:**
Dr Rocio T Martinez Nunez [42].

**Funding acquisition, Leadership and supervision, Metadata curation, Project administration, and Samples and logistics:**
Dr Catherine Ludden [88].

**Funding acquisition, Leadership and supervision, Metadata curation, Samples and logistics, and Sequencing and analysis:**
Dr Sally Corden [69].

**Funding acquisition, Leadership and supervision, Project administration, Samples and logistics, and Sequencing and analysis:**
Ian Johnston [99] and Dr David Bonsall [5].

**Funding acquisition, Leadership and supervision, Sequencing and analysis, Software and analysis tools, and Visualisation:**
Prof Colin P Smith [87] and Dr Ali R Awan [28].

**Funding acquisition, Samples and logistics, Sequencing and analysis, Software and analysis tools, and Visualisation:**
Dr Giselda Bucca [87].

**Leadership and supervision, Metadata curation, Project administration, Samples and logistics, and Sequencing and analysis:**
Dr M. Estee Torok [22, 101].

**Leadership and supervision, Metadata curation, Project administration, Samples and logistics, and Visualisation:**
Dr Kordo Saeed [81, 110] and Dr Jacqui A Prieto [83, 109].

**Leadership and supervision, Metadata curation, Project administration, Sequencing and analysis, and Software and analysis tools:**
Dr David K Jackson [99].

**Metadata curation, Project administration, Samples and logistics, Sequencing and analysis, and Software and analysis tools:**
Dr William L Hamilton [22].

**Metadata curation, Project administration, Samples and logistics, Sequencing and analysis, and Visualisation:**
Dr Luke B Snell [11].

**Funding acquisition, Leadership and supervision, Metadata curation, and Samples and logistics:**
Dr Catherine Moore [69].

**Funding acquisition, Leadership and supervision, Project administration,**and **Samples and logistics:**
Dr Ewan M Harrison [99, 88].

**Leadership and supervision, Metadata curation, Project administration, and Samples and logistics:**
Dr Sonia Goncalves [99].

**Leadership and supervision, Metadata curation, Samples and logistics, and Sequencing and analysis:**
Prof Ian G Goodfellow [24], Dr Derek J Fairley [3, 72], Prof Matthew W Loose [18] and Joanne Watkins [69].

**Leadership and supervision, Metadata curation, Samples and logistics, and Software and analysis tools:**
Rich Livett [99].

**Leadership and supervision, Metadata curation, Samples and logistics, and Visualisation:**
Dr Samuel Moses [25, 106].

**Leadership and supervision, Metadata curation, Sequencing and analysis, and Software and analysis tools:**
Dr Roberto Amato [99], Dr Sam Nicholls [41] and Dr Matthew Bull [69].

**Leadership and supervision, Project administration, Samples and logistics, and Sequencing and analysis:**
Prof Darren L Smith [37, 58, 105].

**Leadership and supervision, Sequencing and analysis, Software and analysis tools, and Visualisation:**
Dr Jeff Barrett [99] and Prof David M Aanensen [14, 114].

**Metadata curation, Project administration, Samples and logistics, and Sequencing and analysis:**
Dr Martin D Curran [65], Dr Surendra Parmar [65], Dr Dinesh Aggarwal [95, 99, 64] and Dr James G Shepherd [48].

**Metadata curation, Project administration, Sequencing and analysis, and Software and analysis tools:**
Dr Matthew D Parker [93].

**Metadata curation, Samples and logistics, Sequencing and analysis, and Visualisation:**
Dr Sharon Glaysher [61].

**Metadata curation, Sequencing and analysis, Software and analysis tools, and Visualisation:**
Dr Matthew Bashton [37, 58], Dr Anthony P Underwood [14, 114], Dr Nicole Pacchiarini [69] and Dr Katie F Loveson [77].

**Project administration, Sequencing and analysis, Software and analysis tools, and Visualisation:**
Dr Alessandro M Carabelli [88].

**Funding acquisition, Leadership and supervision, and Metadata curation:**
Dr Kate E Templeton [53, 90].

**Funding acquisition, Leadership and supervision, and Project administration:**
Dr Cordelia F Langford [99], John Sillitoe [99], Dr Thushan I de Silva [93] and Dr Dennis Wang [93].

**Funding acquisition, Leadership and supervision, and Sequencing and analysis:**
Prof Dominic Kwiatkowski [99, 107], Prof Andrew Rambaut [90], Dr Justin O'Grady [70, 89] and Dr Simon Cottrell [69].

**Leadership and supervision, Metadata curation, and Sequencing and analysis:**
Prof Matthew T.G. Holden [68] and Prof Emma C Thomson [48].

**Leadership and supervision, Project administration, and Samples and logistics:**
Dr Husam Osman [64, 36], Dr Monique Andersson [59], Prof Anoop J Chauhan [61] and Dr Mohammed O Hassan-Ibrahim [6].

**Leadership and supervision, Project administration, and Sequencing and analysis:**
Dr Mara Lawniczak [99].

**Leadership and supervision, Samples and logistics, and Sequencing and analysis:**
Prof Ravi Kumar Gupta [88, 113], Dr Alex Alderton [99], Dr Meera Chand [66], Dr Chrystala Constantinidou [94], Dr Meera Unnikrishnan [94], Prof Alistair C Darby [92], Prof Julian A Hiscox [92] and Prof Steve Paterson [92].

**Leadership and supervision, Sequencing and analysis, and Software and analysis tools:**
Dr Inigo Martincorena [99], Prof David L Robertson [48], Dr Erik M Volz [39], Dr Andrew J Page [70] and Prof Oliver G Pybus [23].

**Leadership and supervision, Sequencing and analysis, and Visualisation:**
Dr Andrew R Bassett [99].

**Metadata curation, Project administration, and Samples and logistics:**
Dr Cristina V Ariani [99], Dr Michael H Spencer Chapman [99, 88], Dr Kathy K Li [48], Dr Rajiv N Shah [48], Dr Natasha G Jesudason [48] and Dr Yusri Taha [50].

**Metadata curation, Project administration, and Sequencing and analysis:**
Martin P McHugh [53] and Dr Rebecca Dewar [53].

**Metadata curation, Samples and logistics, and Sequencing and analysis:**
Dr Aminu S Jahun [24], Dr Claire McMurray [41], Ms Sarojini Pandey [84], Dr James P McKenna [3], Dr Andrew Nelson [58, 105], Dr Gregory R Young [37, 58], Dr Clare M McCann [58, 105] and Mr Scott Elliott [61].

**Metadata curation, Samples and logistics, and Visualisation:**
Ms Hannah Lowe [25].

**Metadata curation, Sequencing and analysis, and Software and analysis tools:**
Dr Ben Temperton [91], Dr Sunando Roy [82], Dr Anna Price [10], Dr Sara Rey [69] and Mr Matthew Wyles [93].

**Metadata curation, Sequencing and analysis, and Visualisation:**
Stefan Rooke [90] and Dr Sharif Shaaban [68].

**Project administration, Samples and logistics, Sequencing and analysis:**
Dr Mariateresa de Cesare [98].

**Project administration, Samples and logistics, and Software and analysis tools:**
Laura Letchford [99].

**Project administration, Samples and logistics, and Visualisation:**
Miss Siona Silveira [81], Dr Emanuela Pelosi [81] and Dr Eleri Wilson-Davies [81].

**Samples and logistics, Sequencing and analysis, and Software and analysis tools:**
Dr Myra Hosmillo [24].

**Sequencing and analysis, Software and analysis tools, and Visualisation:**
Áine O'Toole [90], Dr Andrew R Hesketh [87], Mr Richard Stark [94], Dr Louis du Plessis [23], Dr Chris Ruis [88], Dr Helen Adams [4] and Dr Yann Bourgeois [76].

**Funding acquisition, and Leadership and supervision:**
Dr Stephen L Michell [91], Prof Dimitris Grammatopoulos [84, 112], Dr Jonathan Edgeworth [12], Prof Judith Breuer [30, 82], Prof John A Todd [98] and Dr Christophe Fraser [5].

**Funding acquisition, and Project administration:**
Dr David Buck [98] and Michaela John [9].

**Leadership and supervision, and Metadata curation:**
Dr Gemma L Kay [70].

**Leadership and supervision, and Project administration:**
Steve Palmer [99], Prof Sharon J Peacock [88, 64] and David Heyburn [69].

**Leadership and supervision, and Samples and logistics:**
Danni Weldon [99], Dr Esther Robinson [64, 36], Prof Alan McNally [41, 86], Dr Peter Muir [64], Dr Ian B Vipond [64], Dr John BoYes [29], Dr Venkat Sivaprakasam [46], Dr Tranprit Saluja [75], Dr Samir Dervisevic [54] and Dr Emma J Meader [54].

**Leadership and supervision, and Sequencing and analysis:**
Dr Naomi R Park [99], Karen Oliver [99], Dr Aaron R Jeffries [91], Dr Sascha Ott [94], Dr Ana da Silva Filipe [48], Dr David A Simpson [72] and Dr Chris Williams [69].

**Leadership and supervision, and Visualisation:**
Dr Jane A H Masoli [73, 91].

**Metadata curation, and Samples and logistics:**
Dr Bridget A Knight [73, 91], Dr Christopher R Jones [73, 91], Mr Cherian Koshy [1], Miss Amy Ash [1], Dr Anna Casey [71], Dr Andrew Bosworth [64, 36], Dr Liz Ratcliffe [71], Dr Li Xu-McCrae [36], Miss Hannah M Pymont [64], Ms Stephanie Hutchings [64], Dr Lisa Berry [84], Ms Katie Jones [84], Dr Fenella Halstead [46], Mr Thomas Davis [21], Dr Christopher Holmes [16], Prof Miren Iturriza-Gomara [92], Dr Anita O Lucaci [92], Dr Paul Anthony Randell [38, 104], Dr Alison Cox [38, 104], Pinglawathee Madona [38, 104], Dr Kathryn Ann Harris [30], Dr Julianne Rose Brown [30], Dr Tabitha W Mahungu [74], Dr Dianne Irish-Tavares [74], Dr Tanzina Haque [74], Dr

Jennifer Hart [74], Mr Eric Witele [74], Mrs Melisa Louise Fenton [75], Mr Steven Liggett [79], Dr Clive Graham [56], Ms Emma Swindells [57], Ms Jennifer Collins [50], Mr Gary Eltringham [50], Ms Sharon Campbell [17], Dr Patrick C McClure [97], Dr Gemma Clark [15], Dr Tim J Sloan [60], Mr Carl Jones [15] and Dr Jessica Lynch [2, 111].

**Metadata curation, and Sequencing and analysis:**
Dr Ben Warne [8], Steven Leonard [99], Jillian Durham [99], Dr Thomas Williams [90], Dr Sam T Haldenby [92], Dr Nathaniel Storey [30], Dr Nabil-Fareed Alikhan [70], Dr Nadine Holmes [18], Dr Christopher Moore [18], Mr Matthew Carlile [18], Malorie Perry [69], Dr Noel Craine [69], Prof Ronan A Lyons [80], Miss Angela H Beckett [13], Salman Goudarzi [77], Christopher Fearn [77], Kate Cook [77], Hannah Dent [77] and Hannah Paul [77].

**Metadata curation, and Software and analysis tools:**
Robert Davies [99].

**Project administration, and Samples and logistics:**
Beth Blane [88], Sophia T Girgis [88], Dr Mathew A Beale [99], Katherine L Bellis [99, 88], Matthew J Dorman [99], Eleanor Drury [99], Leanne Kane [99], Sally Kay [99], Dr Samantha McGuigan [99], Dr Rachel Nelson [99], Liam Prestwood [99], Dr Shavanthi Rajatileka [99], Dr Rahul Batra [12], Dr Rachel J Williams [82], Dr Mark Kristiansen [82], Dr Angie Green [98], Miss Anita Justice [59], Dr Adhyana I.K Mahanama [81, 102] and Dr Buddhini Samaraweera [81, 102].

**Project administration, and Sequencing and analysis:**
Dr Nazreen F Hadjirin [88] and Dr Joshua Quick [41].

**Project administration, and Software and analysis tools:**
Mr Radoslaw Poplawski [41].

**Samples and logistics, and Sequencing and analysis:**
Leanne M Kermack [88], Nicola Reynolds [7], Grant Hall [24], Yasmin Chaudhry [24], Malte L Pinckert [24], Dr Iliana Georgana [24], Dr Robin J Moll [99], Dr Alicia Thornton [66], Dr Richard Myers [66], Dr Joanne Stockton [41], Miss Charlotte A Williams [82], Dr Wen C Yew [58], Alexander J Trotter [70], Miss Amy Trebes [98], Mr George MacIntyre-Cockett [98], Alec Birchley [69], Alexander Adams [69], Amy Plimmer [69], Bree Gatica-Wilcox [69], Dr Caoimhe McKerr [69], Ember Hilvers [69], Hannah Jones [69], Dr Hibo Asad [69], Jason Coombes [69], Johnathan M Evans [69], Laia Fina [69], Lauren Gilbert [69], Lee Graham [69], Michelle Cronin [69], Sara Kumziene-SummerhaYes [69], Sarah Taylor [69], Sophie Jones [69], Miss Danielle C Groves [93], Mrs Peijun Zhang [93], Miss Marta Gallis [93] and Miss Stavroula F Louka [93].

**Samples and logistics, and Software and analysis tools:**
Dr Igor Starinskij [48].

**Sequencing and analysis, and Software and analysis tools:**
Dr Chris J Illingworth [47], Dr Chris Jackson [47], Ms Marina Gourtovaia [99], Gerry Tonkin-Hill [99], Kevin Lewis [99], Dr Jaime M Tovar-Corona [99], Dr Keith James [99], Dr Laura Baxter [94], Dr Mohammad T. Alam [94], Dr Richard J Orton [48], Dr Joseph Hughes [48], Dr Sreenu Vattipally [48], Dr Manon Ragonnet-Cronin [39], Dr Fabricia F. Nascimento [39], Mr David Jorgensen [39], Ms Olivia Boyd [39], Ms Lily Geidelberg [39], Dr Alex E Zarebski [23], Dr Jayna Raghwani [23], Dr Moritz UG Kraemer [23], Joel Southgate [10, 69], Dr Benjamin B Lindsey [93] and Mr Timothy M Freeman [93].

**Software and analysis tools, and Visualisation:**
Jon-Paul Keatley [99], Dr Joshua B Singer [48], Leonardo de Oliveira Martins [70], Dr Corin A Yeats [14], Dr Khalil Abudahab [14, 114], Mr Ben EW Taylor [14, 114] and Mirko Menegazzo [14].

**Leadership and supervision:**
Prof John Danesh [99], Wendy Hogsden [46], Dr Sahar Eldirdiri [21], Mrs Anita Kenyon [21], Dr Jenifer Mason [43], Mr Trevor I Robinson [43], Prof Alison Holmes [38, 103], Dr James Price [38, 103], Prof John A Hartley [82], Dr Tanya Curran [3], Dr Alison E Mather [70], Dr Giri Shankar [69], Dr Rachel Jones [69], Dr Robin Howe [69] and Dr Sian Morgan [9].

**Metadata curation:**
Dr Elizabeth Wastenge [53], Dr Michael R Chapman [34, 88, 99], Mr Siddharth Mookerjee [38, 103], Dr Rachael Stanley [54], Mrs Wendy Smith [15], Prof Timothy Peto [59], Dr David Eyre [59], Dr Derrick Crook [59], Dr Gabrielle Vernet [33], Dr Christine Kitchen [10], Huw Gulliver [10], Dr Ian Merrick [10], Prof Martyn Guest [10], Robert Munn [10], Dr Declan T Bradley [63, 72], and Dr Tim Wyatt [63].

**Project administration:**
Dr Charlotte Beaver [99], Luke Foulser [99], Sophie Palmer [88], Carol M Churcher [88], Ellena Brooks [88], Kim S Smith [88], Dr Katerina Galai [88], Georgina M McManus [88], Dr Frances Bolt [38, 103], Dr Francesc Coll [19], Lizzie Meadows [70], Dr Stephen W Attwood [23], Dr Alisha Davies [69], Elen De Lacy [69], Fatima Downing [69], Sue Edwards [69], Dr Garry P Scarlett [76], Mrs Sarah Jeremiah [83] and Dr Nikki Smith [93].

**Samples and logistics:**
Danielle Leek [88], Sushmita Sridhar [88, 99], Sally Forrest [88], Claire Cormie [88], Harmeet K Gill [88], Joana Dias [88], Ellen E Higginson [88], Mailis Maes [88], Jamie Young [88], Michelle Wantoch [7], Sanger Covid Team (www.sanger.ac.uk/covid-team) [99], Dorota Jamrozy [99], Stephanie Lo [99], Dr Minal Patel [99], Verity Hill [90], Ms Claire M Bewshea [91], Prof Sian Ellard [73, 91], Dr Cressida Auckland [73], Dr Ian Harrison [66], Dr Chloe Bishop [66], Dr Vicki Chalker [66], Dr Alex Richter [85], Dr Andrew Beggs [85], Dr Angus Best [86], Dr Benita Percival [86], Dr Jeremy Mirza [86], Dr Oliver Megram [86], Dr Megan Mayhew [86], Dr Liam Crawford [86], Dr Fiona Ashcroft [86], Dr Emma Moles-Garcia [86], Dr Nicola Cumley [86], Mr Richard Hopes [64], Dr Patawee Asamaphan [48], Mr Marc O Niebel [48], Prof Rory N Gunson [100], Dr Amanda Bradley [52], Dr Alasdair Maclean [52], Dr Guy Mollett [52], Dr Rachel Blacow [52], Mr Paul Bird [16], Mr Thomas Helmer [16], Miss Karlie Fallon [16], Dr Julian Tang [16], Dr Antony D Hale [49], Dr Louissa R Macfarlane-Smith [49], Katherine L Harper [49], Miss Holli Carden [49], Dr Nicholas W Machin [45, 64], Ms Kathryn A Jackson [92], Dr Shazaad S Y Ahmad [45, 64], Dr Ryan P George [45], Dr Lance Turtle [92], Mrs Elaine O'Toole [43], Mrs Joanne Watts [43], Mrs Cassie Breen [43], Mrs Angela Cowell [43], Ms Adela Alcolea-Medina [32, 96], Ms Themoula Charalampous [12, 42], Amita Patel [11], Dr Lisa J Levett [35], Dr Judith Heaney [35], Dr Aileen Rowan [39], Prof Graham P Taylor [39], Dr Divya Shah [30], Miss Laura Atkinson [30], Mr Jack CD Lee [30], Mr Adam P Westhorpe [82], Dr Riaz Jannoo [82], Dr Helen L Lowe [82], Miss Angeliki Karamani [82], Miss Leah Ensell [82], Mrs Wendy Chatterton [35], Miss Monika Pusok [35], Mrs Ashok Dadrah [75], Miss Amanda Symmonds [75], Dr Graciela Sluga [44], Dr Zoltan Molnar [72], Mr Paul Baker [79], Prof Stephen Bonner [79], Ms Sarah Essex [79], Dr Edward Barton [56], Ms Debra Padgett [56], Ms Garren Scott [56], Ms Jane Greenaway [57], Dr Brendan AI Payne [50], Dr Shirelle Burton-Fanning [50], Dr Sheila Waugh [50], Dr Veena Raviprakash [17], Ms Nicola Sheriff [17], Ms Victoria Blakey [17], ms Lesley-Anne Williams [17], Dr Jonathan Moore [27], Ms Susanne Stonehouse [27], Dr Louise Smith [55], Dr Rose K Davidson [89], Dr Luke Bedford [26], Dr Lindsay Coupland [54], Ms Victoria Wright [18], Dr Joseph G Chappell [97], Dr Theocharis Tsoleridis [97], Prof Jonathan Ball [97], Mrs Manjinder Khakh [15], Dr Vicki M Fleming [15], Dr Michelle M Lister [15], Dr Hannah C Howson-Wells [15], Dr Louise Berry [15], Dr Tim Boswell [15], Dr Amelia Joseph [15], Dr Iona Willingham [15], Dr Nichola Duckworth [60], Dr Sarah Walsh [60], Dr Emma Wise [2, 111], Dr Nathan Moore [2, 111], Miss Matilde Mori [2, 108, 111], Dr Nick Cortes [2, 111], Dr Stephen Kidd [2, 111], Dr Rebecca Williams [33], Laura Gifford [69], Miss Kelly Bicknell [61], Dr Sarah Wyllie [61], Miss Allyson Lloyd [61], Mr Robert Impey [61], Ms Cassandra S Malone [6], Mr Benjamin J Cogger [6], Nick Levene [62], Lynn Monaghan [62], Dr Alexander J Keeley [93], Dr David G Partridge [78, 93], Dr Mohammad Raza [78, 93], Dr Cariad Evans [78, 93] and Dr Kate Johnson [78, 93].

**Sequencing and analysis:**
Emma Betteridge [99], Ben W Farr [99], Scott Goodwin [99], Dr Michael A Quail [99], Carol Scott [99], Lesley Shirley [99], Scott AJ Thurston [99], Diana Rajan [99], Dr Iraad F Bronner [99], Louise Aigrain [99], Dr Nicholas M Redshaw [99], Dr Stefanie V Lensing [99], Shane McCarthy [99], Alex Makunin [99], Dr Carlos E Balcazar [90], Dr Michael D Gallagher [90], Dr Kathleen A Williamson [90], Thomas D Stanton [90], Ms Michelle L Michelsen [91], Ms Joanna Warwick-Dugdale [91], Dr Robin Manley [91], Ms Audrey Farbos [91], Dr James W Harrison [91], Dr Christine M Sambles [91], Dr David J Studholme [91], Dr Angie Lackenby [66], Dr Tamyo Mbisa [66], Dr Steven Platt [66], Mr Shahjahan Miah [66], Dr David Bibby [66], Dr Carmen Manso [66], Dr Jonathan Hubb [66], Dr Gavin Dabrera [66], Dr Mary Ramsay [66], Dr Daniel Bradshaw [66], Dr Ulf Schaefer [66], Dr Natalie Groves [66], Dr Eileen Gallagher [66], Dr David Lee [66], Dr David Williams [66], Dr Nicholas Ellaby [66], Hassan Hartman [66], Nikos Manesis [66], Vineet Patel [66], Juan Ledesma [67], Ms Katherine A Twohig [67], Dr Elias Allara [64, 88], Ms Clare Pearson [64, 88], Mr Jeffrey K. J. Cheng [94], Dr Hannah E. Bridgewater [94], Ms Lucy R. Frost [94], Ms Grace Taylor-Joyce [94], Dr Paul E Brown [94], Dr Lily Tong [48], Ms Alice Broos [48], Mr Daniel Mair [48], Mrs Jenna Nichols [48], Dr Stephen N Carmichael [48], Dr Katherine L Smollett [40], Dr Kyriaki Nomikou [48], Dr Elihu Aranday-Cortes [48], Ms Natasha Johnson [48], Dr Seema Nickbakhsh [48, 68], Dr Edith E Vamos [92], Dr Margaret Hughes [92], Dr Lucille Rainbow [92], Mr Richard Eccles [92], Ms Charlotte Nelson [92], Dr Mark Whitehead [92], Dr Richard Gregory [92], Mr Matthew Gemmell [92], Ms Claudia Wierzbicki [92], Ms Hermione J Webster [92], Ms Chloe L Fisher [28], Mr Adrian W Signell [20], Dr Gilberto Betancor [20], Mr Harry D Wilson [20], Dr Gaia Nebbia [12], Dr Flavia Flaviani [31], Mr Alberto C Cerda [96], Ms Tammy V Merrill [96], Rebekah E Wilson [96], Mr Marius Cotic [82], Miss Nadua Bayzid [82], Dr Thomas Thompson [72], Dr Erwan Acheson [72], Prof Steven Rushton [51], Prof Sarah O'Brien [51], David J Baker [70], Steven Rudder [70], Alp Aydin [70], Dr Fei Sang [18], Dr Johnny Debebe [18], Dr Sarah Francois [23], Dr Tetyana I Vasylyeva [23], Dr Marina Escalera Zamudio [23], Mr Bernardo Gutierrez [23], Dr Angela Marchbank [10], Joshua Maksimovic [9], Karla Spellman [9], Kathryn McCluggage [9], Dr Mari Morgan [69], Robert Beer [9], Safiah Afifi [9], Trudy Workman [10], William Fuller [10], Catherine Bresner [10], Dr Adrienn Angyal [93], Dr Luke R Green [93], Dr Paul J Parsons [93], Miss Rachel M Tucker [93], Dr Rebecca Brown [93] and Mr Max Whiteley [93].

**Software and analysis tools:**
James Bonfield [99], Dr Christoph Puethe [99], Mr Andrew Whitwham [99], Jennifier Liddle [99], Dr Will Rowe [41], Dr Igor Siveroni [39], Dr Thanh Le-Viet [70] and Amy Gaskin [69].

**Visualisation:**
Dr Rob Johnson [39].

**1** Barking, Havering and Redbridge University Hospitals NHS Trust, **2** Basingstoke Hospital, **3** Belfast Health & Social Care Trust, **4** Betsi Cadwaladr University Health Board, **5** Big Data Institute, Nuffield Department of Medicine, University of Oxford, **6** Brighton and Sussex University Hospitals NHS Trust, **7** Cambridge Stem Cell Institute, University of Cambridge, **8** Cambridge University Hospitals NHS Foundation Trust, **9** Cardiff and Vale University Health Board, **10** Cardiff University, **11** Centre for Clinical Infection & Diagnostics Research, St. Thomas' Hospital and Kings College London, **12** Centre for Clinical Infection and Diagnostics Research, Department of Infectious Diseases, Guy's and St Thomas' NHS Foundation Trust, **13** Centre for Enzyme Innovation, University of Portsmouth (PORT), **14** Centre for Genomic Pathogen Surveillance, University of Oxford, **15** Clinical Microbiology Department, Queens Medical Centre, **16** Clinical Microbiology, University Hospitals of Leicester NHS Trust, **17** County Durham and Darlington NHS Foundation Trust, **18** Deep Seq, School of Life Sciences, Queens Medical Centre, University of Nottingham, **19** Department of Infection Biology, Faculty of Infectious & Tropical Diseases, London School of Hygiene & Tropical Medicine, **20** Department of Infectious Diseases, King's College London, **21** Department of Microbiology, Kettering General Hospital, **22** Departments of Infectious Diseases and Microbiology, Cambridge University Hospitals NHS Foundation Trust; Cambridge, UK, **23** Department of Zoology, University of Oxford, **24** Division of Virology, Department of Pathology, University of Cambridge, **25** East Kent Hospitals University NHS Foundation Trust, **26** East Suffolk and North Essex NHS Foundation Trust, **27** Gateshead Health NHS Foundation Trust, **28** Genomics Innovation Unit, Guy's and St. Thomas' NHS Foundation Trust, **29** Gloucestershire Hospitals NHS Foundation Trust, **30** Great Ormond Street Hospital for Children NHS Foundation Trust, **31** Guy's and St. Thomas' BRC, **32** Guy's and

St. Thomas’ Hospitals, **33** Hampshire Hospitals NHS Foundation Trust, **34** Health Data Research UK Cambridge, **35** Health Services Laboratories, **36** Heartlands Hospital, Birmingham, **37** Hub for Biotechnology in the Built Environment, Northumbria University, **38** Imperial College Hospitals NHS Trust, **39** Imperial College London, **40** Institute of Biodiversity, Animal Health & Comparative Medicine, **41** Institute of Microbiology and Infection, University of Birmingham, **42** King's College London, **43** Liverpool Clinical Laboratories, **44** Maidstone and Tunbridge Wells NHS Trust, **45** Manchester University NHS Foundation Trust, **46** Microbiology Department, Wye Valley NHS Trust, Hereford, **47** MRC Biostatistics Unit, University of Cambridge, **48** MRC-University of Glasgow Centre for Virus Research, **49** National Infection Service, PHE and Leeds Teaching Hospitals Trust, **50** Newcastle Hospitals NHS Foundation Trust, **51** Newcastle University, **52** NHS Greater Glasgow and Clyde, **53** NHS Lothian, **54** Norfolk and Norwich University Hospital, **55** Norfolk County Council, **56** North Cumbria Integrated Care NHS Foundation Trust, **57** North Tees and Hartlepool NHS Foundation Trust, **58** Northumbria University, **59** Oxford University Hospitals NHS Foundation Trust, **60** PathLinks, Northern Lincolnshire & Goole NHS Foundation Trust, **61** Portsmouth Hospitals University NHS Trust, **62** Princess Alexandra Hospital Microbiology Dept., **63** Public Health Agency, **64** Public Health England, **65** Public Health England, Clinical Microbiology and Public Health Laboratory, Cambridge, UK, **66** Public Health England, Colindale, **67** Public Health England, Colindale, **68** Public Health Scotland, **69** Public Health Wales NHS Trust, **70** Quadram Institute Bioscience, **71** Queen Elizabeth Hospital, **72** Queen's University Belfast, **73** Royal Devon and Exeter NHS Foundation Trust, **74** Royal Free NHS Trust, **75** Sandwell and West Birmingham NHS Trust, **76** School of Biological Sciences, University of Portsmouth (PORT), **77** School of Pharmacy and Biomedical Sciences, University of Portsmouth (PORT), **78** Sheffield Teaching Hospitals, **79** South Tees Hospitals NHS Foundation Trust, **80** Swansea University, **81** University Hospitals Southampton NHS Foundation Trust, **82** University College London, **83** University Hospital Southampton NHS Foundation Trust, **84** University Hospitals Coventry and Warwickshire, **85** University of Birmingham, **86** University of Birmingham Turnkey Laboratory, **87** University of Brighton, **88** University of Cambridge, **89** University of East Anglia, **90** University of Edinburgh, **91** University of Exeter, **92** University of Liverpool, **93** University of Sheffield, **94** University of Warwick, **95** University of Cambridge, **96** Viapath, Guy's and St Thomas' NHS Foundation Trust, and King's College Hospital NHS Foundation Trust, **97** Virology, School of Life Sciences, Queens Medical Centre, University of Nottingham, **98** Wellcome Centre for Human Genetics, Nuffield Department of Medicine, University of Oxford, **99** Wellcome Sanger Institute, **100** West of Scotland Specialist Virology Centre, NHS Greater Glasgow and Clyde, **101** Department of Medicine, University of Cambridge, **102** Ministry of Health, Sri Lanka, **103** NIHR Health Protection Research Unit in HCAI and AMR, Imperial College London, **104** North West London Pathology, **105** NU-OMICS, Northumbria University, **106** University of Kent, **107** University of Oxford, **108** University of Southampton, **109** University of Southampton School of Health Sciences, **110** University of Southampton School of Medicine, **111** University of Surrey, **112** Warwick Medical School and Institute of Precision Diagnostics, Pathology, UHCW NHS Trust, **113** Wellcome Africa Health Research Institute Durban and **114** Wellcome Genome Campus.
